# Investigating the effect of sexual behaviour on oropharyngeal cancer risk: a methodological assessment of Mendelian randomization

**DOI:** 10.1101/2021.06.21.21259261

**Authors:** Mark Gormley, Tom Dudding, Linda Kachuri, Kimberley Burrows, Amanda HW Chong, Richard M Martin, Steven Thomas, Jessica Tyrrell, Andrew R Ness, Paul Brennan, Marcus R Munafò, Miranda Pring, Stefania Boccia, Andrew F Olshan, Brenda Diergaarde, Rayjean J Hung, Geoffrey Liu, Eloiza Tajara, Patricia Severino, Tatiana N Toporcov, Martin Lacko, Tim Waterboer, Nicole Brenner, George Davey Smith, Emma E Vincent, Rebecca C Richmond

## Abstract

Human papilloma virus infection is known to influence oropharyngeal cancer (OPC) risk, likely via sexual transmission. However, sexual behaviour has been correlated with other risk factors including smoking and alcohol, meaning independent effects are difficult to establish. Here we evaluate aspects of sexual behaviour in relation to the risk of OPC (2,641 cases and 6,585 controls), using genetic variants associated with age at first sex (AFS) and number of sexual partners (NSP) to perform Mendelian randomization (MR) analyses. While univariable MR showed a causal effect of both later AFS and increasing NSP on OPC, results attenuated in the multivariable models (AFS IVW OR 0.7, 95%CI 0.4, 1.2, *p=* 0.21; NSP IVW OR 0.9, 95%CI 0.5 1.7, *p=* 0.76). We also found evidence for correlated pleiotropy in the genetic instruments for sexual behaviour, emphasising the need for multivariable approaches when performing MR of complex behavioural traits and the triangulation of evidence.

## Introduction

Head and neck squamous cell carcinoma (HNSCC) is a heterogeneous disease ^1^, which can originate from the mucosa of the oral cavity, oropharynx and larynx. Worldwide, there are over half a million incident cases each year, resulting in more than 200,000 deaths annually ^2^. While using tobacco products and consuming alcohol are well-established risk factors across all HNSCC subsites, oral human papilloma virus (HPV) infection has been identified as another risk factor, particularly within the oropharyngeal subsite ^3-6^. In developed countries such as the USA, 60-70% of oropharyngeal cancer (OPC) cases are reported to be HPV- positive ^7^, compared to only around 5% of all oral cancer (OC) cases. Oncogenic HPV type-16 (HPV16) is the most common type found in approximately 90% of HPV-positive oropharyngeal tumours ^8-10^. Antibodies against HPV oncoproteins may be potential biomarkers, with case-control studies demonstrating seropositivity for late (L1) and early (E1, E2, E4, E6, E7) HPV16 proteins were associated with oropharyngeal cancer ^11-14^.

HPV is thought to be sexually transmitted via oro-genital contact ^9,15-20^ and may enter the oropharyngeal mucosa via abrasions in the reticulated tonsillar epithelium ^21^. One large pooled analysis investigating the role of sexual behaviour in HNSCC showed an increased risk of OPC with having a history of six or more lifetime sexual partners (OR 1.3, 95% confidence intervals (95%CI), 1.0, 1.5) and four or more oral sex partners (OR 2.3, 95%CI 1.4, 3.6). A positive association was observed among men who had oral sex (OR 1.6, 95%CI 1.1, 2.3) and those with an earlier age at sexual debut (OR 2.4, 95%CI 1.4, 5.1) ^15^. Conversely, there was no association reported between oral sex practice and head and neck cancer in a more recent meta-analysis of 17 studies (OR 1.1, 95%CI: 0.9, 1.4), suggesting inconsistency in these findings, although 12 of these 17 studies failed to stratify by oral and oropharyngeal subsite ^22^. Furthermore, associations have typically been investigated using case-control studies ^5^, with self-reported sexual behaviour which may be subject to recall bias and misreporting. Positive associations have also been found between sexual behaviour, sexually transmitted infections and other risk factors for HNSCC, including alcohol consumption, indicating the possibility of residual confounding exists ^23^.

Mendelian randomization (MR) is an approach to causal analysis which attempts to overcome shortcomings of conventional observational studies by using single nucleotide polymorphisms (SNPs) which are randomly allocated at conception and known to be reliably associated with modifiable risk factors of interest. These genetic instruments can be used to estimate the effects of risk factors on disease outcomes, in this case sexual behaviours on oropharyngeal cancer ^24,25^, which are less prone to unidentified confounding or reverse causation than conventional epidemiological analysis. Large-scale genome-wide association studies (GWAS) have been performed for sexual behaviour traits, including number of sexual partners (NSP) ^26,27^ and age at first sex (AFS) ^28^. MR makes three key assumptions in that the genetic instrument (i) is robustly associated with the risk factor (i.e., ‘relevance’), (ii) does not share a common cause with the outcome (i.e., ‘exchangeability’), and (iii) affects the outcome only through the risk factor (i.e., ‘exclusion restriction principle’) to check for genetic pleiotropy ^24,25^.

Here, we applied two-sample Mendelian randomization (MR) using summary-level genetic data from the largest available GWAS for each sexual behaviour (sample 1) and oropharyngeal cancer (sample 2). We first conducted univariable MR analysis to assess the effects of NSP and AFS on oropharyngeal cancer risk. We next performed univariable MR analysis to explore the effect of sexual behaviours on HPV seropositivity. Genetic proxies for complex human behaviours are more likely to have broad pleiotropic effects and may influence multiple upstream pathways that indirectly impact on sexual behaviour. In particular, genetic variants associated with sexual behaviour may also influence the disease outcome via other head and neck cancer risk factors, such as smoking and alcohol consumption. For this reason we performed a number of sensitivity analyses: i) MR methods to account for horizontal pleiotropy, ii) MR of sexual behaviours on positive (cervical cancer and seropositivity for Chlamydia trachomatis) and negative control outcomes (lung and oral cancer), iii) Causal Analysis Using Summary Effect estimates (CAUSE), to account for correlated and uncorrelated horizontal pleiotropic effects ^29^, iv) multivariable MR analysis to account for the effects of smoking, alcohol and risk tolerance.

Despite observing an association between genetically predicted AFS and NSP and risk of oropharyngeal cancer using univariable MR, further multivariable analysis indicated violation of the core MR assumptions, likely due to correlated pleiotropy. This highlights the importance of performing comprehensive multivariable MR and sensitivity analyses, in addition to testing for correlated pleiotropy when using genetic instruments to proxy complex human behaviours.

## Results

### Univariable Mendelian Randomization

Using 139 SNPs robustly and independently associated with AFS (**Supplementary Data 1**), there was evidence of a protective effect of later AFS on OPC (IVW OR 0.4, 95%CI 0.3, 0.7, per standard deviation (SD), *p=* <0.001) which was consistent across methods robust to horizontal pleiotropy (MR-Egger, weighted median, and weighted mode) (**Table 1** & **Supplementary Fig.1**). Using 117 SNPs (**Supplementary Data 1**) independently associated with NSP, we found evidence to suggest an adverse effect of increased NSP on the risk of OPC (IVW OR 2.2, 95%CI 1.3, 3.8 per SD, *p=*<0.001). These results were consistent across the other MR methods (**Table 1** & **Supplementary Fig.1**). The protective effect of later AFS was consistent across all geographical regions, with the most precise effects seen in the European (IVW OR 0.4, 95%CI 0.2, 0.8, *p=* <0.001) and North American population (IVW OR 0.4, 95%CI 0.2, 0.8, *p=* 0.01) (**Table 2**). There was also suggestive evidence for an adverse effect of increasing NSP across regions, with the strongest effect again in the North American population (IVW OR 3.0, 95%CI 1.4, 6.5, *p=* 0.01) (**Table 3**).

**Table 1.** Univariable Mendelian randomization results for age at first sex and number of sexual partners on risk of oropharyngeal cancer.

| Outcome | Exposure/<br>Outcome<br>datasets | Outcome<br>N | Controls<br>N | Method | Age at first sex<br>(N SNPs 139) |  | Number of sexual partners<br>(N SNPs 117) |  |
| --- | --- | --- | --- | --- | --- | --- | --- | --- |
|  |  |  |  |  | OR (95%CI) | P | OR (95%CI) | P |
| OPC | UK Biobank/<br>GAME-ON | 2,641 | 6,585 | IVW | 0.44 (0.28, 0.70) | <0.001 | 2.20 (1.27, 3.81) | <0.001 |
| OPC | UK Biobank/<br>GAME-ON | 2,641 | 6,585 | Weighted<br>median | 0.41 (0.23, 0.75) | <0.001 | 2.57 (1.24, 5.29) | 0.01 |
| OPC | UK Biobank/<br>GAME-ON | 2,641 | 6,585 | Weighted<br>mode | 0.23 (0.04, 1.34) | 0.10 | 3.57 (0.58, 21.69) | 0.17 |
| OPC | UK Biobank/<br>GAME-ON | 2,641 | 6,585 | MR-Egger | 0.21 (0.03, 1.37) | 0.10 | 1.88 (0.12, 29.49) | 0.65 |

Abbreviations: OPC, oropharyngeal cancer; IVW, inverse variance weighted; OR, odds ratio; CI, confidence intervals; P, *p*-value; NSP, number of sexual partners; AFS, age at first sex. AFS OR represents the exponential change in odds of oropharyngeal squamous cell carcinoma per SD change (7.3-month delay) in age at first sex. NSP OR represents the exponential change in odds of oropharyngeal squamous cell carcinoma per SD increase (0.94) in number of sexual partners.

**Table 2.** Inverse variance weighted univariable Mendelian randomization results for age at first sex on risk of oropharyngeal cancer, by region.

| Outcome | Region | N SNPs | Outcome N | Control N | Method | OR | CIL | CIU | P value |
| --- | --- | --- | --- | --- | --- | --- | --- | --- | --- |
| Oropharyngeal cancer | Europe | 139 | 1,090 | 2,928 | Inverse variance weighted | 0.36 | 0.17 | 0.78 | <0.001 |
| Oropharyngeal cancer | North America | 139 | 1,119 | 2,329 | Inverse variance weighted | 0.41 | 0.20 | 0.83 | 0.01 |
| Oropharyngeal cancer | South America | 139 | 205 | 727 | Inverse variance weighted | 0.38 | 0.07 | 1.95 | 0.24 |

Abbreviations: SE, standard error; OR, odds ratio; CIL, lower confidence interval; CIU, upper confidence interval; P, *p*-value. OR represents the exponential change in odds of oropharyngeal squamous cell carcinoma per SD change (7.3-month delay) in age at first sex.

**Table 3.**
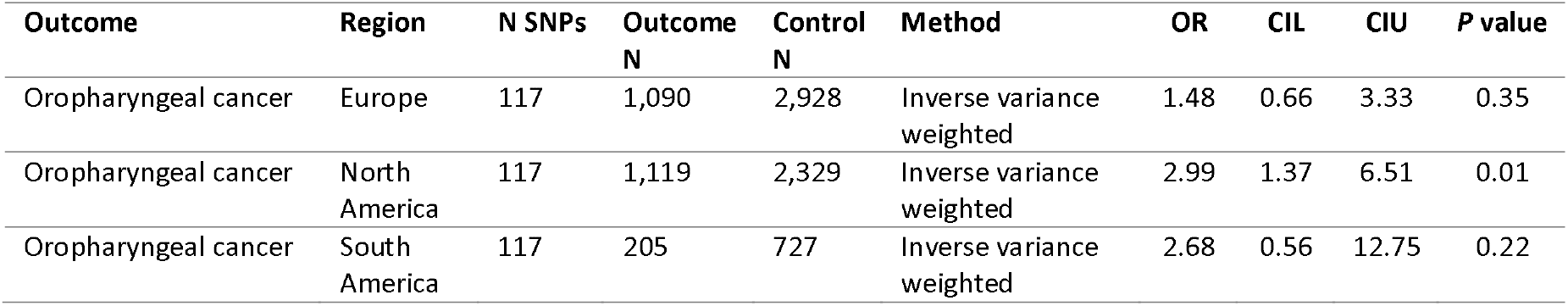
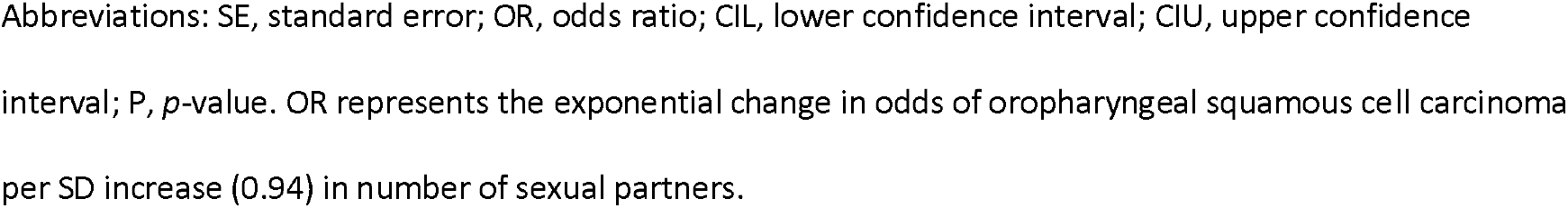
Inverse variance weighted univariable Mendelian randomization results for number of sexual partners on risk of oropharyngeal cancer, by region.

| Outcome | Region | N SNPs | Outcome N | Control N | Method | OR | CIL | CIU | P value |
| --- | --- | --- | --- | --- | --- | --- | --- | --- | --- |
| Oropharyngeal cancer | Europe | 117 | 1,090 | 2,928 | Inverse variance weighted | 1.48 | 0.66 | 3.33 | 0.35 |
| Oropharyngeal cancer | North America | 117 | 1,119 | 2,329 | Inverse variance weighted | 2.99 | 1.37 | 6.51 | 0.01 |
| Oropharyngeal cancer | South America | 117 | 205 | 727 | Inverse variance weighted | 2.68 | 0.56 | 12.75 | 0.22 |

Abbreviations: SE, standard error; OR, odds ratio; CIL, lower confidence interval; CIU, upper confidence interval; P, *p*-value. OR represents the exponential change in odds of oropharyngeal squamous cell carcinoma per SD increase (0.94) in number of sexual partners.

### MR for effect of sexual behaviours on HPV seropositivity

Using the NSP and AFS instruments, we next evaluated the effect of sexual behaviour on the risk of HPV seropositivity in healthy individuals, using a GWAS of serological measures in UK Biobank. There appeared to be some evidence for a protective effect of later AFS (IVW OR 0.5, 95%CI 0.2, 1.0, *p=*0.05) on HPV16 L1 seropositivity (**Table 4** & **Supplementary Table 1**). However, there was limited evidence for a similar protective effect on HPV18 L1, HPV16 E6 or E7 seropositivity. While there was some evidence that increasing NSP also increased the likelihood of HPV16 E6 seropositivity (IVW OR 5.4, 95%CI 1.0, 28.3, *p=*0.05), this was inconsistent among the other tested HPV antibodies (**Table 4** & **Supplementary Table 2**).

**Table 4.**
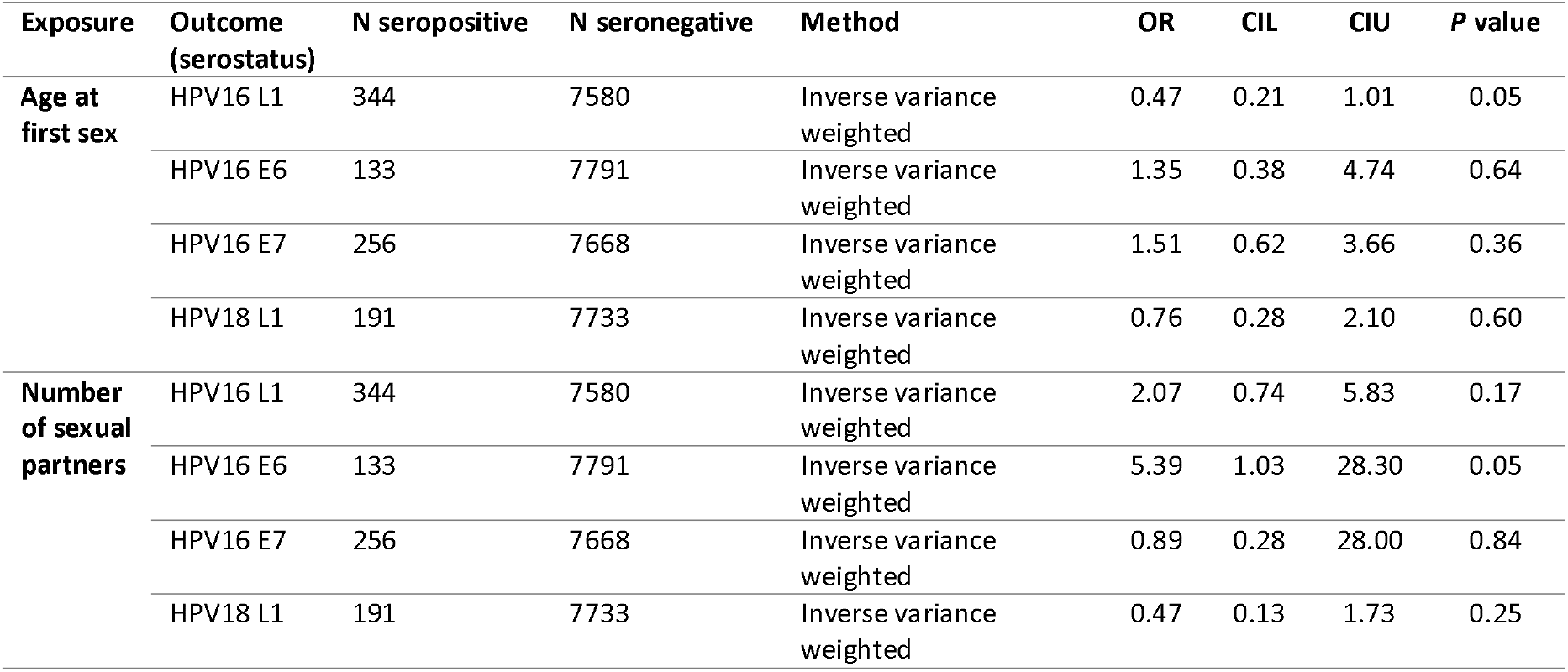
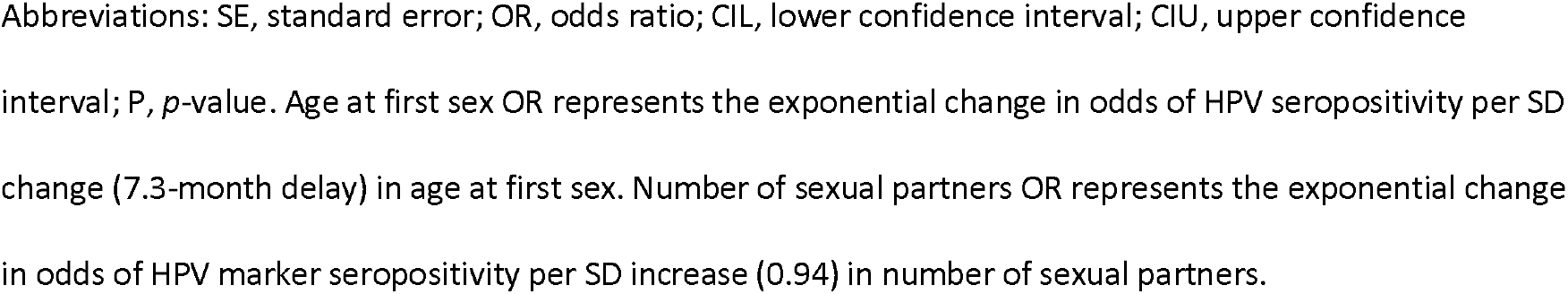
Inverse variance weighted univariable Mendelian randomization results of age at first sex with HPV seropositivity.

| Exposure | Outcome (serostatus) | N seropositive | N seronegative | Method | OR | CIL | CIU | P value |
| --- | --- | --- | --- | --- | --- | --- | --- | --- |
| Age at first sex | HPV16 L1 | 344 | 7580 | Inverse variance weighted | 0.47 | 0.21 | 1.01 | 0.05 |
|  | HPV16 E6 | 133 | 7791 | Inverse variance weighted | 1.35 | 0.38 | 4.74 | 0.64 |
|  | HPV16 E7 | 256 | 7668 | Inverse variance weighted | 1.51 | 0.62 | 3.66 | 0.36 |
|  | HPV18 L1 | 191 | 7733 | Inverse variance weighted | 0.76 | 0.28 | 2.10 | 0.60 |
| Number of sexual partners | HPV16 L1 | 344 | 7580 | Inverse variance weighted | 2.07 | 0.74 | 5.83 | 0.17 |
|  | HPV16 E6 | 133 | 7791 | Inverse variance weighted | 5.39 | 1.03 | 28.30 | 0.05 |
|  | HPV16 E7 | 256 | 7668 | Inverse variance weighted | 0.89 | 0.28 | 28.00 | 0.84 |
|  | HPV18 L1 | 191 | 7733 | Inverse variance weighted | 0.47 | 0.13 | 1.73 | 0.25 |

Abbreviations: SE, standard error; OR, odds ratio; CIL, lower confidence interval; CIU, upper confidence interval; P, *p*-value. Age at first sex OR represents the exponential change in odds of HPV seropositivity per SD change (7.3-month delay) in age at first sex. Number of sexual partners OR represents the exponential change in odds of HPV marker seropositivity per SD increase (0.94) in number of sexual partners.

### Sensitivity analyses

There was limited evidence of weak instrument bias (F-statistic >10) and the proportion of variance in the phenotype (*R*^2^) explained by the genetic instruments ranged from 1 - 2% (**Supplementary Table 3**). There was limited evidence for heterogeneity in the SNP effect estimates for the AFS instrument (QIVW 159.4, *p=* 0.10; Q MR-Egger 158.6, *p=* 0.10), but clear evidence of heterogeneity in the NSP instrument (QIVW 155.6, *p=* 0.007; Q MR-Egger 155.6, *p=* 0.006) (**Supplementary Table 4**). MR-Egger intercepts were not indicative of directional pleiotropy (**Supplementary Table 5**) but there were outliers present on visual inspection in both scatter and leave-one-out plots (**Supplementary Figs.2** & **3**). MR-PRESSO identified 8 outliers for AFS and 7 outliers for NSP, which when corrected for, yielded effects consistent with univariable MR for both instruments (**Supplementary Tables 6-8**). There was evidence of violation of the NOME assumption for both AFS and NSP genetic instruments (i.e., I^2^ statistic <0.90) (**Supplementary Table 9**), so MR-Egger was performed with SIMEX correction. The effects were consistent with previous MR-Egger results for AFS, but there was significant attenuation of the NSP effect on oropharyngeal cancer (SIMEX corrected MR-Egger OR 3.6, 0.4, 32.1, *p=* 0.25) (**Supplementary Table 10**). These estimates should however be interpreted with caution, given evidence of high dilution in the SNP-exposure effects ^30^.

**Table 5.** Multivariable Mendelian randomization for age at first sex with risk of oropharyngeal cancer.

| Exposure | Exposure dataset | N SNPs | F-stat | Q-stat | P-value for instrument validity | Method | AFS OR | 95% CI | P |
| --- | --- | --- | --- | --- | --- | --- | --- | --- | --- |
| Number of Sexual partners | UK Biobank<sup>33</sup> | 152 | 7.81 | 214.14 | 3.77x10<sup>-4</sup> | IVW | 0.42 | 0.19, 0.94 | 0.04 |
|  |  |  |  |  |  | MR Egger | 0.24 | 0.06, 1.01 | 0.05 |
| Comprehensive Smoking Index | UK Biobank<sup>34</sup> | 174 | 8.87 | 191.26 | 0.14 | IVW | 0.48 | 0.25, 0.95 | 0.03 |
|  |  |  |  |  |  | MR-Egger | 0.71 | 0.23, 2.19 | 0.56 |
| Smoking initiation | GSCAN<sup>35</sup> | 215 | 6.43 | 250.58 | 0.04 | IVW | 0.42 | 0.21, 0.83 | 0.01 |
|  |  |  |  |  |  | MR-Egger | 0.61 | 0.21, 1.74 | 0.35 |
| Drinks per week | GSCAN<sup>35</sup> | 147 | 28.88 | 164.77 | 0.11 | IVW | 0.72 | 0.43, 1.20 | 0.21 |
|  |  |  |  |  |  | MR-Egger | 0.43 | 0.14 | 1.28 |
| Risk tolerance | UK Biobank<sup>26</sup> | 160 | 13.68 | 171.18 | 0.21 | IVW | 0.53 | 0.30, 0.93 | 0.03 |
|  |  |  |  |  |  | MR-Egger | 0.24 | 0.08, 0.72 | 0.01 |

Abbreviations: IVW, inverse variance weighted; AFS, age at first sex; OR, odds ratio; CI, confidence intervals; *P*, *p-value*; Q-stat, Cochran's Q statistic; F-stat, conditional F-statistic. AFS OR represents the odds ratio of oropharyngeal squamous cell carcinoma per SD change (7.3-month delay) in age at first sex.

**Table 6.** Multivariable Mendelian randomization for number of sexual partners with risk of oropharyngeal cancer.

| Exposure | Exposure dataset | N SNPs | F-stat | Q-stat | P-value for instrument validity | Method | NSP OR | 95% CI | P |
| --- | --- | --- | --- | --- | --- | --- | --- | --- | --- |
| Age at first sex | UK Biobank<sup>33</sup> | 152 | 7.26 | 214.14 | 3.77x10<sup>-4</sup> | IVW | 0.79 | 0.32, 1.96 | 0.60 |
|  |  |  |  |  |  | MR Egger | 0.97 | 0.35, 2.65 | 0.95 |
| Comprehensive Smoking Index | UK Biobank<sup>34</sup> | 157 | 12.18 | 168.74 | 0.20 | IVW | 0.91 | 0.48, 1.72 | 0.76 |
|  |  |  |  |  |  | MR-Egger | 0.86 | 0.39, 1.88 | 0.70 |
| Smoking initiation | GSCAN<sup>35</sup> | 195 | 7.35 | 204.67 | 0.25 | IVW | 1.51 | 0.77, 2.97 | 0.23 |
|  |  |  |  |  |  | MR-Egger | 1.66 | 0.66, 4.15 | 0.28 |
| Drinks per week | GSCAN<sup>35</sup> | 117 | 22.7 | 151.65 | 0.011 | IVW | 1.45 | 0.75, 2.79 | 0.27 |
|  |  |  |  |  |  | MR-Egger | 1.61 | 0.76, 3.41 | 0.21 |
| Risk tolerance | UK Biobank<sup>26</sup> | 125 | 7.06 | 145.56 | 0.072 | IVW | 2.04 | 0.93, 4.44 | 0.07 |
|  |  |  |  |  |  | MR-Egger | 2.12 | 0.66, 6.83 | 0.21 |

Abbreviations: IVW, inverse variance weighted; NSP, number of sexual partners; OR, odds ratio; CI, confidence intervals; *P*, *p-value*; Q-stat, Cochran's Q statistic; F-stat, conditional F-statistic. NSP OR represents the odds ratio of oropharyngeal squamous cell carcinoma per SD increase (0.94) in number of sexual partners.

### Positive and negative control analyses

Univariable MR analysis conducted within UK Biobank found a protective effect for later AFS on cervical cancer, which is known to be another HPV-driven cancer type (IVW OR 0.4, 95%CI 0.3, 0.7, *p=* <0.001) (**Supplementary Table 11**). A similar effect was found when assessing the effect of AFS on C. trachomatis seropositivity based on pGP3 antigen, another positive control (IVW OR 0.4, 95%CI 0.3, 0.6, *p=* <0.001) (**Supplementary Table 11**). There was also evidence for an adverse effect of increasing NSP on cervical cancer risk (IVW OR 1.9, 95CI% 1.0, 3.9, *p=* 0.06) and a positive association between NSP and C. trachomatis serostatus (IVW OR 2.4, 95%CI 1.4, 4.1, *p=* <0.001) (**Supplementary Table 12**).

Using lung cancer as a negative control, in univariable MR there was a strong protective effect of AFS (IVW OR 0.1 95%CI, 0.1, 0.3 *p=* <0.001) (**Supplementary Table 11**) and an adverse effect of increasing NSP (IVW OR 7.1 95%CI, 2.4, 21.6 *p=* <0.001) (**Supplementary Table 12**), indicating violation of the MR assumptions. A protective effect was also observed in relation to AFS with oral cancer, another negative control (IVW OR 0.6, 95%CI 0.4, 1.0, *p=* 0.03) (**Supplementary Table 11**); however, there was no effect for NSP on oral cancer (IVW OR 1.2, 95%CI 0.7, 2.0, *p=* 0.47) (**Supplementary Table 12**).

While there was no strong evidence for directional pleiotropy (**Supplementary Table 13**), there was some evidence of heterogeneity (**Supplementary Table 14**) for both AFS and NSP in the lung and oral cancer analyses, suggesting that pleiotropy may be present ^31^. While scatter and leave-one-out plots showed no obvious outliers (**Supplementary Figs.4-7**), MR-PRESSO identified outliers for AFS and for NSP across all positive and negative controls.

When corrected for outliers, the lung cancer results remained consistent with the univariable MR, suggesting further violation of the MR assumptions for the AFS and NSP instruments (**Supplementary Tables 15-17**).

### Investigating correlated pleiotropy using CAUSE

We used GWAS summary statistics to evaluate evidence for an effect of AFS and NSP on oropharyngeal cancer, using the Causal Analysis using Summary Effect estimates (CAUSE) method to account for correlated pleiotropy ^32^. For AFS, CAUSE suggested there was relatively similar evidence for sharing (correlated pleiotropy) (*p=* 0.02) and causal models (*p=* 0.05) compared to the null (no effect) model (**Supplementary Table 18** & **Supplementary Fig.8**). Comparing both shared and causal models, there was limited evidence that the causal model fit the data better than the sharing model (*p=* 0.44), indicating that correlated pleiotropy could not be discounted. When investigating the causal effect of NSP on oropharyngeal cancer, neither shared (*p=* 0.30) nor causal (*p=* 0.27) models appeared to fit in comparison to the null model, providing limited evidence for a causal effect of NSP (**Supplementary Table 19** & **Supplementary Fig.9**).

### Multivariable MR

In total there were 21 overlapping SNPs identified between genetic instruments (**Supplementary Table 20**) and LD score regression highlighted strong genetic correlation between the traits (*rg* = |0.20-0.63|) **Supplementary Table 21** & **Supplementary Fig.10**). Multivariable MR analysis was therefore carried out to investigate the direct causal effect of AFS and NSP on oropharyngeal cancer after accounting for the other sexual behaviour, smoking, alcohol, and risk tolerance. While the effect of NSP diminished (IVW OR 0.8, 95%CI 0.3, 2.0, *p=* 0.60), the AFS effect remained (IVW OR 0.4, 95%CI 0.2, 0.9, *p=* 0.04), after accounting for the other sexual behaviour in multivariable MR (**Tables 5** & **6; Fig.1**). When accounting for smoking and risk tolerance, the effect of AFS remained consistent within the oropharyngeal subsite (**Table 5** & **Fig.2**). However, there was attenuation of the effect for AFS when controlling for drinks per week (IVW OR 0.7, 95%CI 0.4, 1.2, *p=* 0.21). The effect of NSP on oropharyngeal cancer attenuated when accounting for lifetime smoking (IVW OR 0.9, 95%CI 0.5 1.72, *p=* 0.76), alcohol consumption (IVW OR 1.5, 95%CI 0.8, 2.8, *p=* 0.27) and risk tolerance (IVW OR 2.0, 95%CI 0.9, 4.4, *p=* 0.07) (**Table 6** & **Fig.3**) These results suggest the NSP and AFS instruments may include pleiotropic variants related to smoking and drinking behaviours. Some of the multivariable models including smoking initiation and drinks per week showed high levels of heterogeneity and therefore further risk of invalid instruments (**Tables 5** & **6**). However, the MR-Egger intercepts in the multivariable analyses were consistent with the null, indicative of no further directional pleiotropy (**Supplementary Table 22**) and the effects estimated were also consistent across both IVW and MR-Egger models (**Tables 5** & **6**).

**Fig.1.**
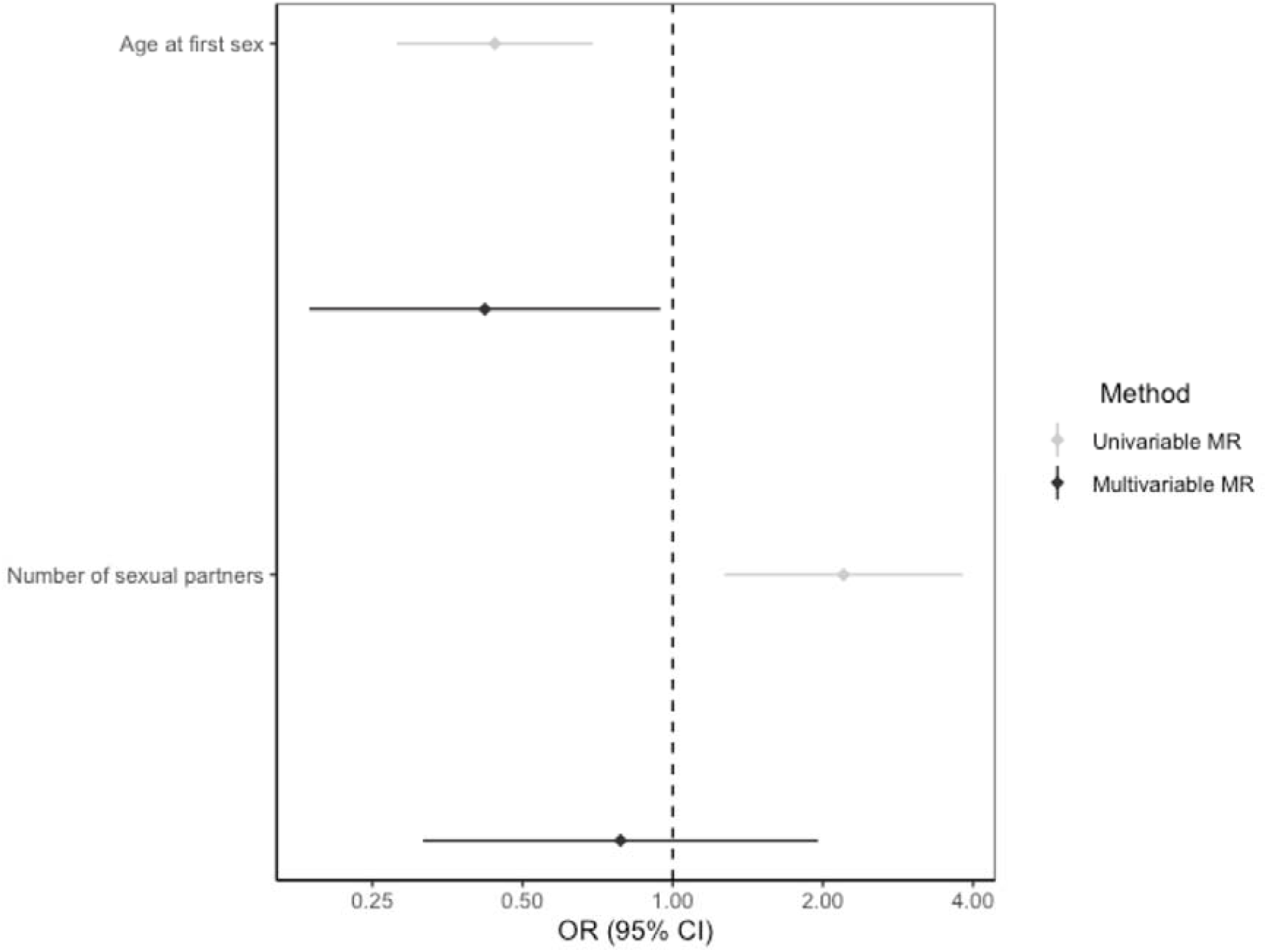
Forest plot comparing univariable and multivariable Mendelian randomization effects of age at first sex and number of sexual partners on oropharyngeal cancer risk. Effect estimates are reported on the log odds scale with 95% confidence intervals. Age at first sex point estimate represents the exponential change in odds of oropharyngeal squamous cell carcinoma per SD change (7.3-month delay) in age at first sex. Number of sexual partners point estimate represents the exponential change in odds of oropharyngeal squamous cell carcinoma per SD increase (0.94) in number of sexual partners.

**Fig.2.**
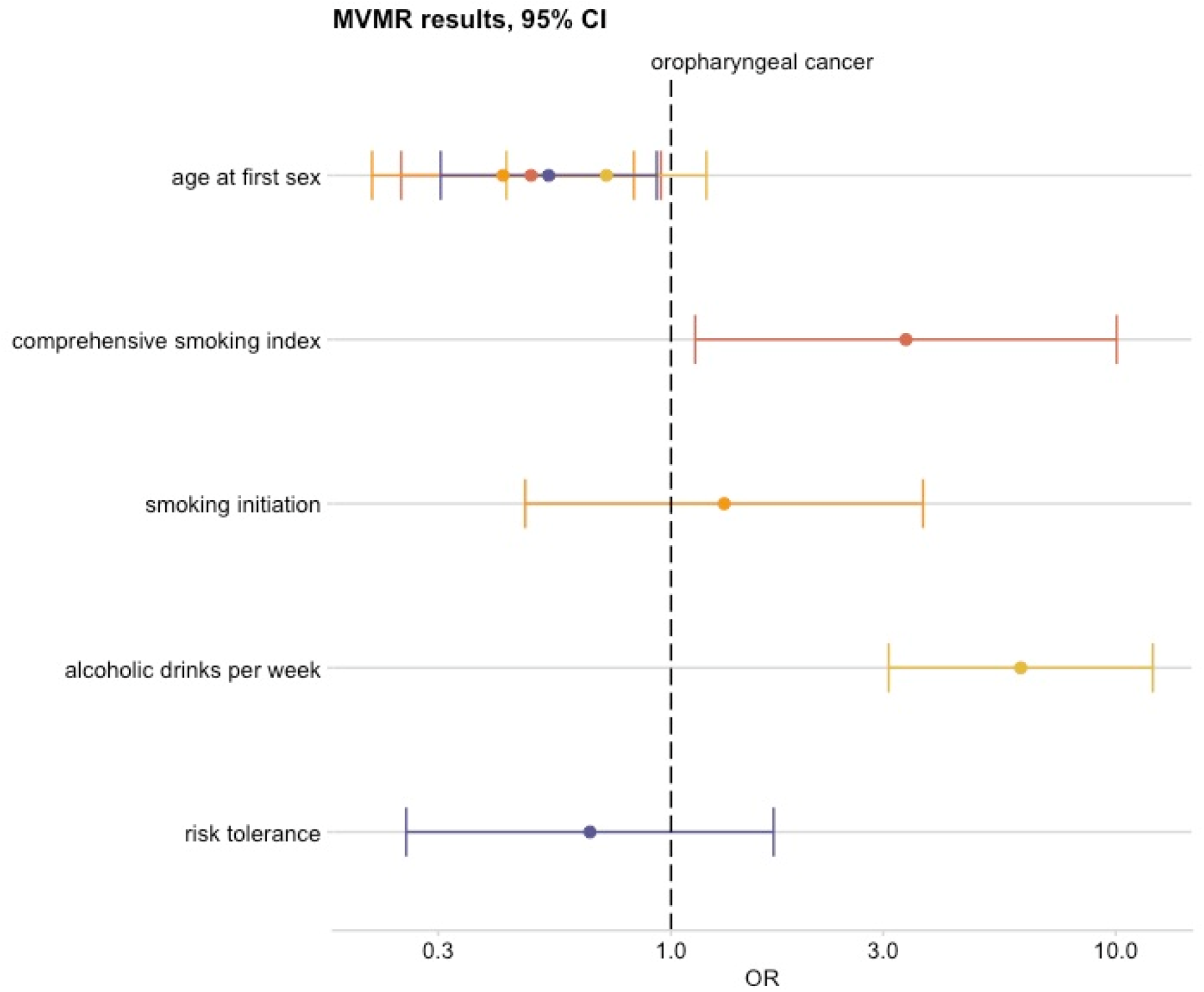
Forest plot showing multivariable Mendelian randomization results for age at first sex single nucleotide polymorphisms with risk of oropharyngeal cancer. Effect estimates on oropharyngeal cancer risk are reported on the log odds scale with 95% confidence intervals. Age at first sex OR represents the change in odds of oropharyngeal squamous cell carcinoma per SD change (7.3-month delay) in age at first sex. Comprehensive smoking index (dark orange), smoking initiation (light orange), alcoholic drinks per week (yellow), risk tolerance (blue).

**Fig.3.**
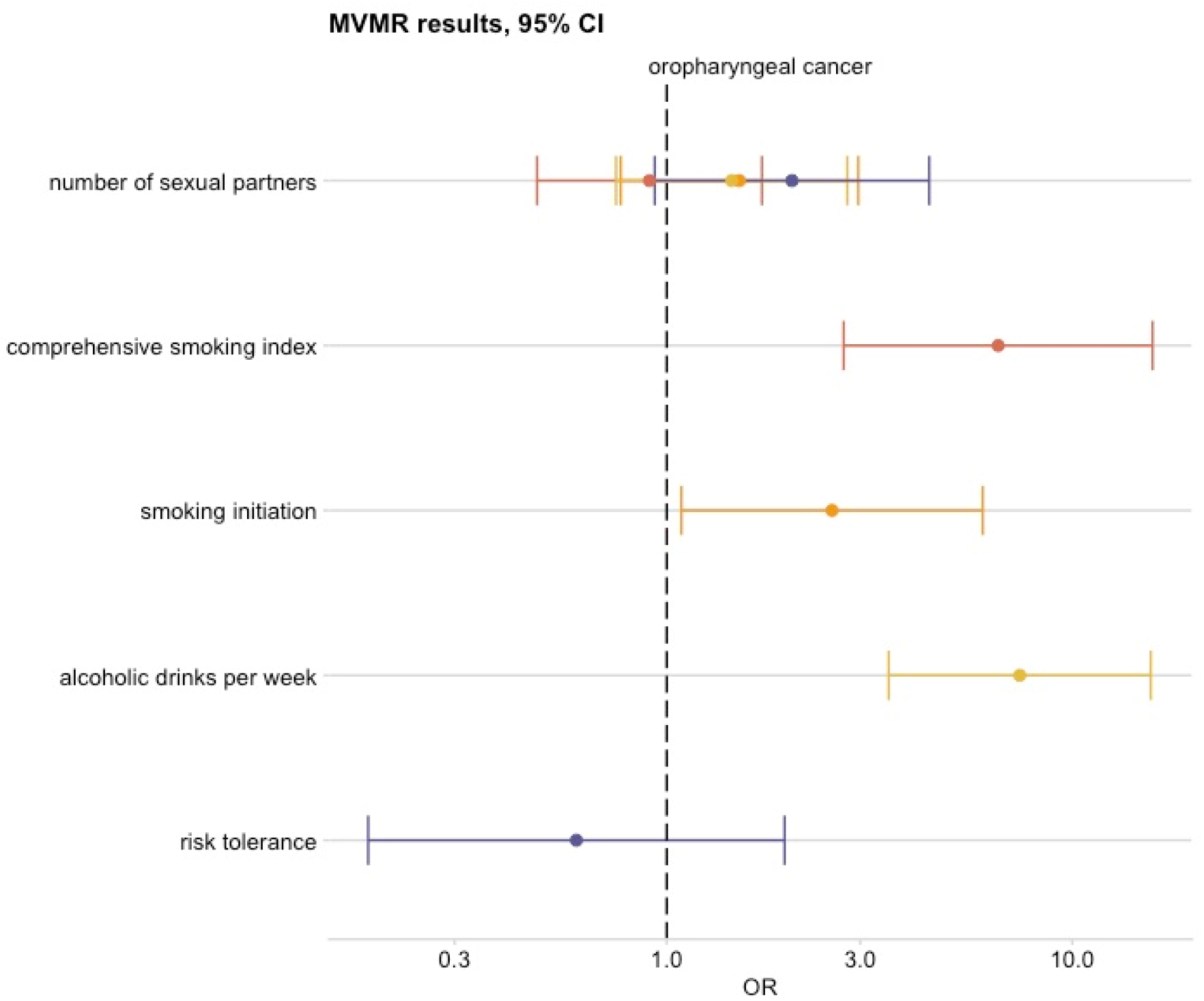
Forest plot showing multivariable Mendelian randomization results for number of sexual partners single nucleotide polymorphisms with risk of oropharyngeal cancer. Effect estimates on oropharyngeal cancer risk are reported on the log odds scale with 95% confidence intervals. Number of sexual partners OR represents the change in odds of oropharyngeal squamous cell carcinoma per SD change (0.94) in number of sexual partners. Comprehensive smoking index (dark orange), smoking initiation (light orange), alcoholic drinks per week (yellow), risk tolerance (blue).

In additional multivariable MR analysis of AFS and NSP on lung cancer, effects for both instruments were attenuated once smoking was included in the model. With AFS, this was clearly seen when controlling for smoking initiation (IVW OR 1.1, 95%CI 0.8, 1.6, *p=* 0.57) and a change in direction of the effect of AFS was evident when controlling for the comprehensive smoking index (IVW OR 2.0, 95%CI 1.3, 3.0, p<0.001) (**Supplementary Table 23** & **Supplementary Fig.11**). Similarly, there was limited evidence for an effect of NSP on lung cancer when controlling for the comprehensive smoking index (IVW OR 0.7, 95%CI 0.4, 1.1, *p=* 0.09). The MR-Egger intercept deviated from the null in the multivariable models including smoking, suggestive of further directional pleiotropy in this analysis (**Supplementary Table 24**).

## Discussion

In this study we applied Mendelian randomization to evaluate the effects of both later age at first sex and increased number of sexual partners on the risk of oropharyngeal cancer. We observed convergence between genetic pathways influencing sexual behaviors and susceptibility to oropharyngeal cancer, which may be partly mediated by HPV infection, however, we also uncovered complex correlated pleiotropy with other putative risk factors. Univariable MR results suggested a protective effect of later age at first sex and an adverse effect of increased number of sexual partners. However, these effects attenuated in the multivariable MR analyses that controlled for smoking behaviour and alcohol consumption. While there was suggestive evidence for an effect of sexual behaviours on some HPV16 serology measures and in cervical cancer (supportive of a causal mechanism via HPV infection), the same direction of effect was observed in negative control analysis (lung and oral cancer) indicating potential violation of the MR assumptions. Furthermore, CAUSE provided less support for a causal effect of AFS and NSP on oropharyngeal cancer risk, highlighting the risk of correlated pleiotropy in the genetic instruments for these complex behavioural traits.

### Sexual behaviours and HPV transmission

Over 90% of HPV positive OPC is caused by the high-risk genotype 16, with almost all oral infections thought to be sexually acquired ^36^. HPV is a small non-enveloped DNA virus, with its genome encoding for both early oncoproteins E6/E7 and the late capsid proteins such as L1. The overexpression of these oncogenes is thought to stimulate proliferation and lateral expansion of epithelial basal cells, progressing to a malignant phenotype. HPV E6 forms a complex which leads to rapid degradation of tumour suppressor protein p53, resulting in deregulation of cell cycle checkpoints. E7 binds to a complex which ubiquitinates another tumour suppressor protein, retinoblastoma (pRb), again resulting in uncontrolled G1/S phase of the cell cycle ^37^. While the transmission of HPV via sexual intercourse is well known and HPV in turn a major risk factor for cervical malignancies, the role of HPV in oropharyngeal cancer risk has only been acknowledged in recent decades ^8^. Among OPC cases, HPV16 E6 serology is a good biomarker (∼99% specificity, >90% sensitivity) and therefore both E6 and E7 are highly associated with this disease ^38^. However, when studying these antibodies in the general population, E6 seroprevalence appears to be very low (0.5- 1%), but in comparison with low incidence rates of HPV-positive OPC this figure is still high, suggesting that not all individuals in the general population who have HPV16 E6 seropositivity will develop an oropharyngeal tumour or other HPV-associated cancer ^38^. Consequently, we performed this analysis in UK Biobank and observed a strong and consistent association with sexual behavior. In our univariable MR analysis, the effects of AFS and NSP instruments on risk of HPV16 and HPV18 seropositivity were not consistent, compared with recent observational studies which demonstrate an association between serology markers and sexual behaviour responses in UK Biobank ^38^. This could be as a result of the small number of seropositive HPV16 (*n*= <450) and HPV18 (*n*= 265) cases within the UK Biobank pilot study used in our genetic analysis or that results from genetic proxies and questionnaire data are not directly comparable ^39^. Using serology measures to predict HPV seropositivity or a HPV-positive OPC diagnosis is not straightforward, often requiring the use of multiple markers simultaneously ^40^. Going forward, more reliable tests may emerge which could improve our prediction of both the infection and disease.

### Regional differences in sexual behaviour and HPV prevalence

Although the incidence of OPC in South America is similar to that in Western Europe and North America, the prevalence of HPV16 is reportedly low ^41^. Latin America has an estimated overall HPV-positive head and neck cancer prevalence of between 3 – 4%, compared with 25% in European and North American populations ^41-43^. This could partly be explained by differences in data collection and methods used to detect HPV. Despite Latin American countries having an average age of sexual debut between 18-19 years old ^44^, the International Head and Neck Cancer Epidemiology (INHANCE) Consortium found that these countries reported higher mean numbers of sexual partners (e.g., Brazil n=22), compared with North American (e.g., USA, Atlanta n=10) or European (e.g., Warsaw *n*= 15) populations ^15^. Stratifying by region in our univariable MR analysis, we found a consistent protective effect for AFS and similarly, a consistent increased risk effect for NSP across all three regions (Europe, North America, and South America), with evidence for the most precise effects in the North American population. In the largest pooled analysis, authors also report possible recall or reporting biases, given that some of the sexual behaviour interviews were carried out with family members nearby, in addition to small sample sizes (<150 cases) ^15^ which may have affected their results.

### Confounding by other risk factors

While transmission of HPV to the upper aerodigestive tract is thought to be through oral sexual contact ^9,15-20^, a more recent meta-analysis reported no association between oral sex practices and head and neck cancer risk ^22^. This could be explained by the inclusion of older studies ^22^, which may not have captured the more recent rise in number of HPV-positive OPC cases which has been described by some as an ‘epidemic’ and predicted to overtake oral cancer within the next decade ^45^. However, a study in the UK found that there was no change in the proportion of HPV-attributable cases from 2002-2011, although the incidence of OPC doubled over the same time period and national surveys have not described an increase in oral sex behaviour ^1,46^. In one multi-national study of 1,626 men aged 18–73 years with 4-year follow-up, no association was detected between oral sexual behaviours and incident HPV infection, but oral oncogenic HPV was found to be more prevalent in current smokers compared with non-smokers ^47^. Furthermore, tobacco exposure induces proinflammatory and immunosuppressive effects, which could potentially increase the likelihood of HPV infection and persistence ^48,49^. Since risk factors such as smoking and alcohol consumption are strongly associated with sexual behaviour and are well established in the aetiology of HNSCC, this may confound the relationship between sexual behaviours with HPV transmission and similarly oropharyngeal cancer in observational studies ^50,51^.

Although Mendelian randomization analysis minimises the likelihood of confounding, since germline genetic variants should not theoretically be influenced by subsequent environmental confounders, pleiotropy is a major concern whereby genetic variants associated with the exposure (sexual behaviours) are related to the outcome (oropharyngeal cancer) through alternative, independent biological pathways. We used a series of analyses to evaluate the potential for pleiotropy. We first performed several methods (MR-Egger ^52^, weighted median ^53^ and weighted mode ^54^) which allow for the existence of horizontal pleiotropy and correct for this. We also identified and corrected for outlier SNPs most likely to exhibit pleiotropic effects. In univariable MR analyses, estimates were consistent with an effect of AFS and NSP on oropharyngeal cancer risk. However, in further MR analysis taking lung cancer as a negative control, we observed the same direction of effect for AFS and NSP which we did not expect, since there is no plausible biological mechanism directly linking sexual behaviour with lung cancer risk. Evidence of an effect here indicates potential violation of the MR assumptions.

Strong genetic correlation between sexual behaviours and other risk factors such as smoking, alcohol and general risk tolerance were found using LD score regression. The genetic instruments used in MR may therefore comprise variants which primarily influence other risk factors, which could induce correlated pleiotropy (**Fig.4**). We conducted two subsequent analyses to evaluate this. The CAUSE approach provided limited evidence for any effect of NSP on oropharyngeal cancer and was unable to distinguish an effect of AFS from the situation of correlated pleiotropy. We also performed multivariable MR to control for alcohol, smoking and risk tolerance, so as to determine the direct causal effect of sexual behaviours on oropharyngeal cancer. Effect estimates attenuated when alcohol and smoking were taken into account in the multivariable MR models, again highlighting the role of potential pleiotropy in the genetic instruments for sexual behaviour.

**Fig.4.**
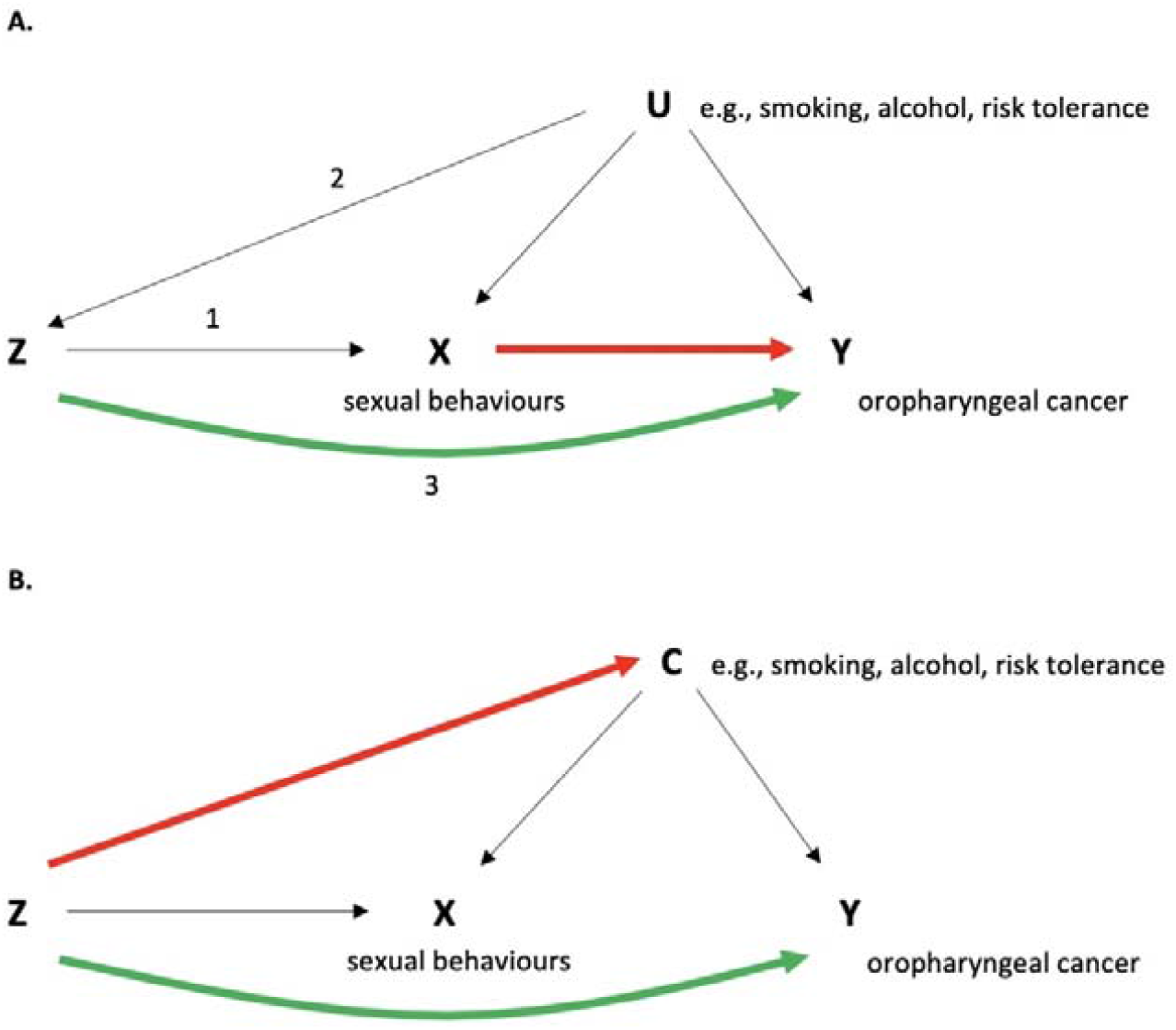
**A**. Genetic variants (Z) act as proxies or instruments to investigate if an exposure (X), is associated with a disease outcome (Y). Causal inference can be made between X and Y if the following conditions are upheld: (1) Z is a valid instrument, reliably associated with X (‘relevance’); (2) Z is independent of any measured or unmeasured confounding factors (U) (‘exchangeability’) and (3) there is no independent association between Z and Y except through X (‘exclusion restriction’). **B.** Directed acyclic graph (DAG) depicting correlated pleiotropy (C) whereby the genetic variant (Z) can affect the exposure (X) and the outcome (Y) via a shared heritable factor (C), for example here through smoking, alcohol, or general risk tolerance.

### Strengths and limitations

MR was employed in this study in an attempt to overcome the drawbacks of conventional epidemiological studies. However, MR makes various assumptions which if violated may generate spurious conclusions. For example, sexual behaviours are difficult to instrument genetically due to measurement error (e.g., as a result of reporting bias) and because they are time-varying as well as context and culture-dependent. This could hamper the detection of genetic associations related to these traits which has implications for genetic instrument strength (the first assumption of MR), given the low percentage of variation explained (*R*^2^), as well as potential violation of the no measurement error (NOME) assumption, with relatively low I^2^ values.

Additionally, the available genetic instruments are not specifically for oral sex, which is the conceptually relevant exposure and mode of HPV transmission. However, other sexual behaviours are likely to be correlated and developing genetic instruments for specific sexual activities poses some methodological and ethical challenges. While the random inheritance of genetic variants from parents to offspring means genotypes are typically much less associated with many potential confounders than directly measured exposures (the second MR assumption), an obvious violation of this is created due to population stratification which can introduce confounding of genotype-outcome associations. Although the GWAS for both NSP and AFS were adjusted for genetic principal components, given that sexual behaviours are strongly socially patterned, residual population structure may reintroduce confounding into MR analysis. Although a rare outcome, there is potential sample overlap present as head and neck cancer cases were not excluded from previously published AFS or NSP GWAS, however recent studies suggest the incurred bias is much less substantial than that due to weak instruments, or overestimation of the SNP-trait effect ^55,56^.

The third major assumption of MR is the exclusion restriction principle (i.e., that the genetic variant affects the outcome exclusively through its effect on the exposure). We performed a series of comprehensive sensitivity analyses to evaluate potential violation of this assumption. While several pleiotropy-robust (MR-Egger, weighted median, and weighted mode) and outlier exclusion methods provided limited evidence for violation of this assumption, the results of the lung cancer negative control analysis, CAUSE method and multivariable MR all suggested violation of the exclusion restriction assumption in the univariable MR of sexual behaviours on oropharyngeal cancer risk. When multiple sources of evidence provide conflicting estimates, it is necessary to appraise the relative biases of the approaches in order to best “triangulate” evidence ^57,58^. In this instance, it is possible that the primary phenotype for the genetic variants used to instrument the sexual behaviours has been mis-specified. For example, the genetic variants may be primarily associated with other traits (e.g., risk taking) and indirectly to sexual behaviours via the primary traits. Similarly, sexual behaviour instruments may be associated with traits which don’t have a direct negative connotation. In this instance, the Instrument Strength independent of Direct Effect (InSIDE) assumption of approaches such as MR-Egger is likely to be violated, whereas the CAUSE is less vulnerable to environmental confounders that are correlated with genetic variants than the other pleiotropy-robust methods. Multivariable MR was also used to directly model the potential indirect effects of the genetic variants via other traits (smoking, alcohol, and risk tolerance) and supported the conclusions of the CAUSE method. Finally, we could not distinguish between HPV positive and negative oropharyngeal tumours in the GAME-ON summary data, which would require further analysis at an independent level or a GWAS of oropharyngeal cancer stratified by HPV status. The GWAS-by-subtraction approach ^59^ could be useful to account for latent factors of other behavioral traits to identify more specific genetic instruments for sexual behaviour, if valid instruments for these traits exist. More serological data may become available in the UK Biobank and other clinical genetic studies, which could enhance power to evaluate potential the extent to which any effect of sexual behaviour on cancer risk is mediated by HPV.

## Conclusions

In conclusion, this study used a comprehensive series of MR analyses to investigate sexual behaviours in relation to oropharyngeal cancer. We initially observed an association between genetically predicted AFS and NSP and risk of oropharyngeal cancer using univariable MR. Despite using genetic variants strongly related to these traits in large-scale GWAS, further multivariate methods indicated violation of the core MR assumptions, likely due to correlated pleiotropy. Effect estimates attenuated when alcohol and smoking were taken into account in the multivariable MR models, highlighting the importance of performing these further analyses, particularly when using genetic instruments which proxy complex behavioural traits.

## Methods

### Summary-level data for sexual behaviours

Summary statistics for AFS were obtained from a GWAS conducted in the UK Biobank (n=397,338) ^28^. AFS was treated as a continuous variable, with individuals considered as eligible if they had given a valid answer to the question *“What was your age when you first had sexual intercourse? (Sexual intercourse includes vaginal, oral or anal intercourse)”* and ages <12 years old were excluded. Since AFS had a non-normal distribution, a within-sex inverse rank normal transformation was applied ^28^. Where possible the full 272 SNP AFS instrument was used, except in the primary analysis of OPC, whereby only 139 SNPs could be extracted from head and neck cancer data. We obtained summary statistics for the NSP instrument (117 SNPs) from a GWAS conducted in UK Biobank ^26^ (n=⍰370,711). NSP was treated as a continuous variable based on responses to the question: *“About how many sexual partners have you had in your lifetime?”*. Respondents who reported >99 lifetime sexual partners were asked to confirm their responses and a value of zero was assigned to participants who reported having never had sex, which was normalised separately for both males and females with an inverse rank normal transformation ^26^. Both AFS and NSP GWAS adjusted for the top 10 principal components (accounting for population stratification), sex, and birth year. For AFS, those participants with family data were controlled with non-independence of family members or else one family member was included in the analysis ^28^.

### Summary-level data for oropharyngeal cancer

The largest available GWAS for oropharyngeal cancer was performed on 2,641 OPC cases and 6,585 matched controls from 12 studies which were part of the Genetic Associations and Mechanisms in Oncology (GAME-ON) Network ^60^. Cancer cases comprised the following ICD-10 codes: oropharynx (C01.9, C02.4 and C09.0–C10.9). Stratification was conducted by geographical region to evaluate potential heterogeneity in any effects given potential differences in the distribution of genetic variants for specific traits within populations. As GAME-ON included participants from Europe (45.3%), North America (43.9%) and South America (10.8%), this study was restricted to individuals of predominantly European ancestry to avoid the effect of population structure. Details of the studies included as well as the genotyping and imputation performed have been described previously ^60,61^.

### Univariable Mendelian randomization

To assess effects of NSP and AFS, we used SNPs which reached genome-wide significance (p <5×10^−8^) in the respective GWAS and for which pairwise r^2^ <0.1 (with 250kb linkage disequilibrium (LD) windows), ensuring only independent SNPs were selected into the instrument. Two-sample MR analyses were conducted using the “TwoSampleMR” package (version 0.5.5) in R (version 4.0.2) to extract the SNPs instrumenting the risk factor from the oropharyngeal cancer GWAS. Harmonization of the direction of effects between exposure and outcome associations was performed and palindromic SNPs were aligned when minor allele frequencies (MAFs) were less than 0.3 or were otherwise excluded. SNP specific Wald estimates were calculated (SNP-outcome estimate divided by SNP-exposure estimate) and an inverse variance weighted (IVW) method applied to meta-analyse these in order to obtain an effect estimate of the risk factor on oropharyngeal cancer risk.

### MR for sexual behaviours on HPV and C. trachomatis seropositivity

Where there was evidence for an effect of sexual behaviour on oropharyngeal cancer risk, we also aimed to confirm the suspected aetiological link via HPV, by investigating the effects of NSP and AFS on a range of seropositivity measures against HPV16 L1, E6, E7 and HPV18 L1 proteins. Here, seropositivity suggests previous HPV exposure, which can be a predictor of cancer. Generally, HPV16 L1 antibodies are considered cumulative exposure markers, while HPV16 E6 and E7 have been associated with HPV-driven cancers but not all those who test positive are expected to develop a HPV-driven cancer ^38^. Summary-level genetic data for HPV16 and HPV18 serological measures were obtained from UK Biobank. We performed individual GWAS for each measure using a similar approach as described by Kachuri et al. ^62^. Details on how these GWAS were conducted can be found in the **Supplementary information**.

### Sensitivity analyses

The strength of each genetic instrument was determined by the magnitude and precision of association with the sexual behaviour, which was considered to be sufficient if the corresponding F-statistic was >10. The fixed-effect IVW method provides an unbiased estimate in the absence of horizontal pleiotropy or when horizontal pleiotropy is balanced ^31^. To account for directional pleiotropy, we compared results with three other MR methods, which each make different assumptions about this: MR-Egger ^52^, weighted median ^53^ and weighted mode ^54^. Further detail on these methods is provided in the **Supplementary information (see “Methods”)**.

### Positive and negative control analyses

To further assess the specificity and sensitivity of the genetic instruments identified in relation to sexual behaviour, we conducted additional positive and negative control MR analyses. These aimed to appraise the role of AFS and NSP on a) cervical cancer and C. trachomatis seropositivity, as positive control outcomes where evidence of an effect would support the aetiological link via HPV; and b) lung cancer and oral cancer as negative controls, where a direct causal effect of sexual behaviour is unlikely and so where any evidence of an effect would indicate potential violation of the MR assumptions due to pleiotropy, population stratification or selection bias ^63^. Details on the GWAS summary data used to conduct positive and negative control outcomes can be found in the Supplementary information.

### Causal Analysis using Summary Effect estimates (CAUSE)

While sensitivity analyses like MR-Egger, weighted median and weighted mode can detect horizontal or uncorrelated pleiotropy, whereby the genetic variant affects the exposure (sexual behaviours) and outcome (oropharyngeal cancer) through separate mechanisms, correlated pleiotropy is an alternative scenario which could generate spurious associations in MR. Here, the genetic variant affects the exposure and outcome via a shared heritable factor. Correlated pleiotropy may be present in the genetic instruments for AFS and NSP, which if undetected could lead to false positive results (**Fig.4**).

We used the CAUSE method in an attempt to identify potential correlated pleiotropy ^29^. CAUSE proposes that any causal effect of an exposure on the outcome leads to correlation for all variants with a non-zero effect on the exposure, while a shared factor induces correlation for only a subset of exposure effect variants ^29^. GWAS summary statistics were used to generate two models nested in a “null” effects model. The sharing model allows for horizontal pleiotropic effects but no causal effect (γ = 0), whereas the causal model has γ as a free parameter. The Bayesian expected log pointwise posterior density (ELPD) is used to compare models, producing a one-sided p value which tests the best fitting model. In particular, if the hypothesis that the sharing model fits the data at least as well as the causal model is rejected, we can conclude that the data are consistent with a causal effect ^29^.

### LD Score regression

Genetic correlation was calculated between the two sexual behaviour traits (AFS and NSP), smoking, alcohol, and general risk tolerance. Summary-level genome-wide association studies were obtained for alcohol consumption (drinks per week, *n*= 941,280) and smoking initiation (a binary phenotype indicating whether an individual had ever smoked regularly) (*n*= 1,232,091) from the GSCAN meta-analysis ^35^. Summary statistics were also obtained from a GWAS of general risk tolerance (n=⍰939,908), derived from a meta-analysis of UK Biobank (n=⍰431,126) question *“Would you describe yourself as someone who takes risks?”* and 23andMe (n=⍰508,782) question *“Overall, do you feel comfortable or uncomfortable taking risks?”*. The GWAS of risk tolerance was based on one’s tendency or willingness to take risks, making them more likely to engage in risk-taking behaviours more generally ^26^. The regression was performed using pre-computed LD scores calculated based on individuals of European ancestry from 1000 Genomes European data and are appropriate for use with European-ancestry GWAS data ^64^. This was filtered to HapMap3 SNPs as these are well-imputed in most studies ^65^. SNPs found on chromosome 6 in the region 26MB to 34MB were excluded. GWAS summary statistics were converted for LD score regression using the munge_sumstats.py command from the command line tool “ldsc”, and LD score regression was performed using the ldsc.py command.

### Multivariable Mendelian randomization

To account for the potential genetic overlap with other risk factors ^26^ for oropharyngeal cancer which may lead to correlated pleiotropy, we next conducted two-sample multivariable MR analysis. This accounted for the effects of the other sexual behaviour, smoking, alcohol consumption and risk tolerance in the MR of each sexual behaviour onto the cancer outcomes. First multivariable MR was carried out to assess the effect of genetic overlap between AFS and NSP using the genome-wide significant SNPs identified as instruments in the univariable analysis (272 SNPs for AFS and 117 SNPs for NSP). 196 independent SNPs (p <5 ×10^−8^) were used in the analysis for smoking initiation, 60 SNPs for alcoholic drinks per week and 123 for risk tolerance, after excluding SNPs with a pairwise r^2^ > 0.001. To better capture lifetime smoking (duration, heaviness, and cessation), we used 108 SNPs which make up the comprehensive smoking index, derived by Wootton et al in the UK Biobank (*n*= 462,690) ^34^. SNP overlap was assessed between all instruments. We used generalised versions of Cochran’s Q statistical tests for both instrument strength and validity^66^. Both the IVW and MR-Egger framework have been extended to estimate causal effects in multivariable MR analysis ^67,68^, which was conducted using both the MVMR (version 0.2.0) and MendelianRandomization ^69^ (version 0.5.0) packages in R (version 4.0.2)

Causal Analysis using Summary Effect Estimates, LD Score Regression and Multivariable Mendelian randomization approaches all require full GWAS summary data for the proposed risk factors of interested. Where full data were available for the GWAS of NSP ^26^, these have yet to be published for the GWAS of AFS, which we used to select genetic instruments. Therefore, for these approaches, we used another GWAS for AFS, also conducted using UK Biobank data (n=406,457), for which full summary data are publicly available (https://gwas.mrcieu.ac.uk/datasets/ukb-b-6591/). This GWAS was conducted using the MRC IEU UK Biobank GWAS pipeline, more details of which can be found in Elsworth et al, 2019 ^70^.

## Supporting information

Conflict of interest

## Data Availability

Data availability
Summary-level analysis was conducted using publicly available GWAS data. Full summary statistics for the GAME-ON GWAS can be accessed via dbGAP (OncoArray: Oral and Pharynx Cancer; study accession number: phs001202.v1.p1) and via the IEU OpenGWAS project https://gwas.mrcieu.ac.uk/. Access to UK Biobank (https://www.ukbiobank.ac.uk/) data is available to researchers through application. For the purpose of open access, the author has applied a CC BY public copyright licence to any Author Accepted Manuscript version arising from this submission. UK Biobank approval was given for this project (ID 40644 Investigating aetiology, associations and causality in diseases of the head and neck) and UK Biobank GWAS data was also accessed under the application (ID 15825 MR-Base: an online resource for Mendelian randomization using summary data-Dr Philip Haycock).
Code availability statement
MR analyses were conducted using the TwoSampleMR package in R (version 3.5.3). A copy of the code and all files used in this analysis is available at: https://github.com/rcrichmond/sexual_behaviours_opc

## Acknowledgements

M.G. was a National Institute for Health Research (NIHR) academic clinical fellow and is currently supported by a Wellcome Trust GW4-Clinical Academic Training PhD Fellowship. This research was funded in part, by the Wellcome Trust [Grant number 220530/Z/20/Z]. For the purpose of open access, the author has applied a CC BY public copyright licence to any Author Accepted Manuscript version arising from this submission. R.C.R. is a de Pass VC research fellow at the University of Bristol. J.T. is supported by an Academy of Medical Sciences (AMS) Springboard award, which is supported by the AMS, the Wellcome Trust, Global Challenges Research Fund (GCRF), the Government Department of Business, Energy and Industrial strategy, the British Heart Foundation and Diabetes UK (SBF004\1079). R.M.M. was supported by a Cancer Research UK (C18281/A20919) programme grant (the Integrative Cancer Epidemiology Programme). R.M.M. and A.R.N. are supported by the National Institute for Health Research (NIHR) Bristol Biomedical Research Centre which is funded by the National Institute for Health Research (NIHR) and is a partnership between University Hospitals Bristol NHS Foundation Trust and the University of Bristol. Department of Health and Social Care disclaimer: The views expressed are those of the authors and not necessarily those of the NHS, the NIHR or the Department of Health and Social Care. This publication presents data from the Head and Neck 5000 study. The study was a component of independent research funded by the National Institute for Health Research (NIHR) under its Programme Grants for Applied Research scheme (RP-PG-0707-10034). The views expressed in this publication are those of the author(s) and not necessarily those of the NHS, the NIHR or the Department of Health. Core funding was also provided through awards from Above and Beyond, University Hospitals Bristol and Weston Research Capability Funding and the NIHR Senior Investigator award to A.R.N. Human papillomavirus (HPV) serology was supported by a Cancer Research UK Programme Grant, the Integrative Cancer Epidemiology Programme (C18281/A20919). B.D. and the University of Pittsburgh head and neck cancer case-control study are supported by US National Institutes of Health (NIH) grants: P50 CA097190, P30 CA047904 and R01 DE025712. The genotyping of the HNSCC cases and controls was performed at the Center for Inherited Disease Research (CIDR) and funded by the US National Institute of Dental and Craniofacial Research (NIDCR; 1X01HG007780-0). The University of North Carolina (UNC) CHANCE study was supported in part by the National Cancer Institute (R01-CA90731). E.E.V is supported by Diabetes UK (17/0005587). E.E.V is also supported by the World Cancer Research Fund (WCRF UK), as part of the World Cancer Research Fund International grant programme (IIG_2019_2009). E.H.T and P.S. were supported by FAPESP grant 10/51168-0 (GENCAPO/Head and Neck Genome project). M.G., T.D., K.B., A.C., R.M.M., M.M., G.D.S, E.E.V. and R.C.R are part of the Medical Research Council Integrative Epidemiology Unit at the University of Bristol supported by the Medical Research Council (MC_UU_00011/1, MC_UU_00011/5, MC_UU_00011/6, MC_UU_00011/7).

## Author Contributions

M.G. and R.C.R. conceived the study and M.G. carried out data curation and analysis, validating the results separately. L.K. completed both the HPV and cervical cancer GWAS and helped with interpretation of these data. T.W. and N.B. produced serology data for HPV in the UK Biobank pilot and provided expertise on interpretation of these data. Head and neck cancer summary genetic data was obtained through multiple collaborations from studies lead by A.R.N., S.T., A.F.O., R.J.H., G.L., B.D., S.B., E.T., P.S., T.N.T., M.L. and P.B. The initial manuscript was drafted by M.G., L.K., G.D.S. and R.C.R. Expert guidance on MR methodology was provided by L.K., T.D., K.B., B.D., R.C.R., G.D.S, R.M.M. All authors M.G., T.D., L.K., K.B., A.C., R.M.M., S.T., J.T., A.R.N., P.B., M.M., M.P., S.B., A.F.O., B.D., R.J.H., G.L., E.T., P.S., T.N.T., M.L., T.W., N.B., G.D.S., E.V. and R.C.R. contributed to the interpretation of the results and critical revision of the manuscript. M.G. supervisory team includes R.C.R., E.V., J.T., A.R.N and G.D.S.

## Data availability

Summary-level analysis was conducted using publicly available GWAS data. Full summary statistics for the GAME-ON GWAS can be accessed via dbGAP (OncoArray: Oral and Pharynx Cancer; study accession number: phs001202.v1.p1) and via the IEU OpenGWAS project https://gwas.mrcieu.ac.uk/. Access to UK Biobank (https://www.ukbiobank.ac.uk/) data is available to researchers through application. For the purpose of open access, the author has applied a CC BY public copyright licence to any Author Accepted Manuscript version arising from this submission. UK Biobank approval was given for this project (ID 40644 “Investigating aetiology, associations and causality in diseases of the head and neck”) and UK Biobank GWAS data was also accessed under the application (ID 15825 “MR-Base: an online resource for Mendelian randomization using summary data”- Dr Philip Haycock).”

## Code availability statement

MR analyses were conducted using the “TwoSampleMR” package in R (version 3.5.3). A copy of the code and all files used in this analysis is available at: https://github.com/rcrichmond/sexual_behaviours_opc

## References

1 Thomas, S. J., Penfold, C. M., Waylen, A. & Ness, A. R. The changing aetiology of head and neck squamous cell cancer: A tale of three cancers? Clin Otolaryngol 43, 999–1003 (2018).

2 Sung, H. et al. Global cancer statistics 2020: GLOBOCAN estimates of incidence and mortality worldwide for 36 cancers in 185 countries. CA: A Cancer Journal for Clinicians (2021).

3 Syrjanen, K., Syrjanen, S., Lamberg, M., Pyrhonen, S. & Nuutinen, J. Morphological and immunohistochemical evidence suggesting human papillomavirus (HPV) involvement in oral squamous cell carcinogenesis. Int J Oral Surg 12, 418–424 (1983).

4 Smith, E. M. et al. Human papillomavirus seropositivity and risks of head and neck cancer. Int J Cancer 120, 825–832 (2007).

5 D’Souza, G. et al. Case-control study of human papillomavirus and oropharyngeal cancer. New Engl J Med 356, 1944–1956 (2007).

6 Pan, C., Issaeva, N. & Yarbrough, W. G. HPV-driven oropharyngeal cancer: current knowledge of molecular biology and mechanisms of carcinogenesis. Cancers Head Neck 3, 12 (2018).

7 Chaturvedi, A. K. et al. Worldwide Trends in Incidence Rates for Oral Cavity and Oropharyngeal Cancers. Journal of Clinical Oncology 31, 4550–4559 (2013).

8 Gillison, M. L. et al. Evidence for a causal association between human papillomavirus and a subset of head and neck cancers. J Natl Cancer Inst 92, 709–720 (2000).

9 Gillison, M. L., Chaturvedi, A. K., Anderson, W. F. & Fakhry, C. Epidemiology of Human Papillomavirus-Positive Head and Neck Squamous Cell Carcinoma. J Clin Oncol 33, 3235–3242 (2015).

10 Castellsagué, X. et al. HPV Involvement in Head and Neck Cancers: Comprehensive Assessment of Biomarkers in 3680 Patients. J Natl Cancer Inst 108, djv403 (2016).

11 Kreimer, A. R. et al. Evaluation of human papillomavirus antibodies and risk of subsequent head and neck cancer. Journal of clinical oncology : official journal of the American Society of Clinical Oncology 31, 2708–2715 (2013).

12 Kreimer, A. R. et al. Kinetics of the Human Papillomavirus Type 16 E6 Antibody Response Prior to Oropharyngeal Cancer. JNCI: Journal of the National Cancer Institute 109 (2017).

13 Anantharaman, D. et al. Human papillomavirus infections and upper aero-digestive tract cancers: the ARCAGE study. J Natl Cancer Inst 105, 536–545 (2013).

14 Ribeiro, K. B. et al. Low human papillomavirus prevalence in head and neck cancer: results from two large case-control studies in high-incidence regions. Int J Epidemiol 40, 489–502 (2011).

15 Heck, J. E. et al. Sexual behaviours and the risk of head and neck cancers: a pooled analysis in the International Head and Neck Cancer Epidemiology (INHANCE) consortium. Int J Epidemiol 39, 166–181 (2010).

16 Herrero, R. et al. Human papillomavirus and oral cancer: the International Agency for Research on Cancer multicenter study. J Natl Cancer Inst 95, 1772–1783 (2003).

17 Schwartz, S. M. et al. Oral cancer risk in relation to sexual history and evidence of human papillomavirus infection. J Natl Cancer Inst 90, 1626–1636 (1998).

18 Rajkumar, T. et al. Oral cancer in Southern India: the influence of body size, diet, infections and sexual practices. Eur J Cancer Prev 12, 135–143 (2003).

19 Smith, E. M. et al. Age, sexual behavior and human papillomavirus infection in oral cavity and oropharyngeal cancers. Int J Cancer 108, 766–772 (2004).

20 Shah, A. et al. Oral sex and human papilloma virus-related head and neck squamous cell cancer: a review of the literature. Postgrad Med J 93, 704–709 (2017).

21 Doorbar, J. & Griffin, H. Refining our understanding of cervical neoplasia and its cellular origins. Papillomavirus Res 7, 176–179 (2019).

22 Farsi, N. J. et al. Sexual behaviours and head and neck cancer: A systematic review and meta-analysis. Cancer Epidemiol 39, 1036–1046 (2015).

23 Khadr, S. N. et al. Investigating the relationship between substance use and sexual behaviour in young people in Britain: findings from a national probability survey. BMJ Open 6, e011961 (2016).

24 Davey Smith, G. & Hemani, G. Mendelian randomization: genetic anchors for causal inference in epidemiological studies. Hum Mol Genet 23, R89–98 (2014).

25 Smith, G. D. & Ebrahim, S. ‘Mendelian randomization’: can genetic epidemiology contribute to understanding environmental determinants of disease? Int J Epidemiol 32, 1–22 (2003).

26 Karlsson Linner, R. et al. Genome-wide association analyses of risk tolerance and risky behaviors in over 1 million individuals identify hundreds of loci and shared genetic influences. Nat Genet 51, 245–257 (2019).

27 Ganna, A. et al. Large-scale GWAS reveals insights into the genetic architecture of same-sex sexual behavior. Science 365, eaat7693 (2019).

28 Mills, M. C. et al. Identification of 370 loci for age at onset of sexual and reproductive behaviour, highlighting common aetiology with reproductive biology, externalizing behaviour and longevity. bioRxiv, 2020.2005.2006.081273 (2020).

29 Morrison, J., Knoblauch, N., Marcus, J. H., Stephens, M. & He, X. Mendelian randomization accounting for correlated and uncorrelated pleiotropic effects using genome-wide summary statistics. Nat Genet (2020).

30 Bowden, J. et al. Assessing the suitability of summary data for two-sample Mendelian randomization analyses using MR-Egger regression: the role of the I2 statistic. Int J Epidemiol 45, 1961–1974 (2016).

31 Hemani, G., Bowden, J. & Smith, G. D. Evaluating the potential role of pleiotropy in Mendelian randomization studies. Hum Mol Genet 27, R195–R208 (2018).

32 Morrison, J., Knoblauch, N., Marcus, J. H., Stephens, M. & He, X. Mendelian randomization accounting for correlated and uncorrelated pleiotropic effects using genome-wide summary statistics. Nat Genet 52, 740–747 (2020).

33 Elsworth, B. et al. The MRC IEU OpenGWAS data infrastructure. bioRxiv, 2020.2008.2010.244293 (2020).

34 Wootton, R. E. et al. Evidence for causal effects of lifetime smoking on risk for depression and schizophrenia: a Mendelian randomisation study. Psychol Med, 1–9 (2019).

35 Liu, M. Z. et al. Association studies of up to 1.2 million individuals yield new insights into the genetic etiology of tobacco and alcohol use. Nat Genet 51, 237-+ (2019).

36 Pan, C., Issaeva, N. & Yarbrough, W. G. HPV-driven oropharyngeal cancer: current knowledge of molecular biology and mechanisms of carcinogenesis. Cancers of the Head & Neck 3, 12 (2018).

37 Chung, C. H. & Gillison, M. L. Human Papillomavirus in Head and Neck Cancer: Its Role in Pathogenesis and Clinical Implications. Clinical Cancer Research 15, 6758–6762 (2009).

38 Brenner, N. et al. Characterization of human papillomavirus (HPV) 16 E6 seropositive individuals without HPV-associated malignancies after 10 years of follow-up in the UK Biobank. EBioMedicine 62, 103123 (2020).

39 Mentzer, A. J. et al. Identification of host-pathogen-disease relationships using a scalable Multiplex Serology platform in UK Biobank. medRxiv, 19004960 (2019).

40 Dahlstrom, K. R. et al. HPV Serum Antibodies as Predictors of Survival and Disease Progression in Patients with HPV-Positive Squamous Cell Carcinoma of the Oropharynx. Clinical cancer research : an official journal of the American Association for Cancer Research 21, 2861–2869 (2015).

41 Perdomo, S., Martin Roa, G., Brennan, P., Forman, D. & Sierra, M. S. Head and neck cancer burden and preventive measures in Central and South America. Cancer Epidemiology 44, S43–S52 (2016).

42 Kreimer, A. R., Clifford, G. M., Boyle, P. & Franceschi, S. Human papillomavirus types in head and neck squamous cell carcinomas worldwide: a systematic review. Cancer Epidemiol Biomarkers Prev 14, 467–475 (2005).

43 Dayyani, F. et al. Meta-analysis of the impact of human papillomavirus (HPV) on cancer risk and overall survival in head and neck squamous cell carcinomas (HNSCC). Head Neck Oncol 2, 15 (2010).

44 Gayet, C., Juarez, F. & Bozon, M. Vol. 5 67–90 (2013).

45 Bosetti, C. et al. Global trends in oral and pharyngeal cancer incidence and mortality. Int J Cancer 147, 1040–1049 (2020).

46 Schache, A. G. et al. HPV-Related Oropharynx Cancer in the United Kingdom: An Evolution in the Understanding of Disease Etiology. Cancer Res 76, 6598–6606 (2016).

47 Kreimer, A. R. et al. Incidence and clearance of oral human papillomavirus infection in men: the HIM cohort study. Lancet (London, England) 382, 877–887 (2013).

48 Castle, P. E. How does tobacco smoke contribute to cervical carcinogenesis? J Virol 82, 6084–6086 (2008).

49 Arnson, Y., Shoenfeld, Y. & Amital, H. Effects of tobacco smoke on immunity, inflammation and autoimmunity. J Autoimmun 34, J258–265 (2010).

50 Hashibe, M. et al. Interaction between Tobacco and Alcohol Use and the Risk of Head and Neck Cancer: Pooled Analysis in the International Head and Neck Cancer Epidemiology Consortium. Cancer Epidem Biomar 18, 541–550 (2009).

51 Gormley, M. et al. A multivariable Mendelian randomization analysis investigating smoking and alcohol consumption in oral and oropharyngeal cancer. Nat Commun 11, 6071 (2020).

52 Bowden, J., Davey Smith, G. & Burgess, S. Mendelian randomization with invalid instruments: effect estimation and bias detection through Egger regression. Int J Epidemiol 44, 512–525 (2015).

53 Bowden, J., Smith, G. D., Haycock, P. C. & Burgess, S. Consistent Estimation in Mendelian Randomization with Some Invalid Instruments Using a Weighted Median Estimator. Genet Epidemiol 40, 304–314 (2016).

54 Hartwig, F. P., Smith, G. D. & Bowden, J. Robust inference in summary data Mendelian randomization via the zero modal pleiotropy assumption. Int J Epidemiol 46, 1985–1998 (2017).

55 Mounier, N. & Kutalik, Z. Correction for sample overlap, winner’s curse and weak instrument bias in two-sample Mendelian Randomization. bioRxiv, 2021.2003.2026.437168 (2021).

56 Minelli, C. et al. The use of two-sample methods for Mendelian randomization analyses on single large datasets. bioRxiv, 2020.2005.2007.082206 (2020).

57 Lawlor, D. A., Tilling, K. & Davey Smith, G. Triangulation in aetiological epidemiology. Int J Epidemiol 45, 1866–1886 (2017).

58 Munafo, M. R. & Davey Smith, G. Robust research needs many lines of evidence. Nature 553, 399–401 (2018).

59 Demange, P. A. et al. Investigating the genetic architecture of noncognitive skills using GWAS-by-subtraction. Nat Genet 53, 35–44 (2021).

60 Lesseur, C. et al. Genome-wide association analyses identify new susceptibility loci for oral cavity and pharyngeal cancer. Nat Genet 48, 1544–1550 (2016).

61 Dudding, T. et al. Assessing the causal association between 25-hydroxyvitamin D and the risk of oral and oropharyngeal cancer using Mendelian randomization. Int J Cancer 143, 1029–1036 (2018).

62 Kachuri, L. et al. The landscape of host genetic factors involved in immune response to common viral infections. Genome Medicine 12, 93 (2020).

63 Sanderson, E., Richardson, T. G., Hemani, G. & Davey Smith, G. The use of negative control outcomes in Mendelian randomization to detect potential population stratification. Int J Epidemiol (2021).

64 Auton, A. et al. A global reference for human genetic variation. Nature 526, 68–74 (2015).

65 Altshuler, D. M. et al. Integrating common and rare genetic variation in diverse human populations. Nature 467, 52–58 (2010).

66 Sanderson, E., Davey Smith, G., Windmeijer, F. & Bowden, J. An examination of multivariable Mendelian randomization in the single-sample and two-sample summary data settings. Int J Epidemiol (2018).

67 Rees, J. M. B., Wood, A. M. & Burgess, S. Extending the MR-Egger method for multivariable Mendelian randomization to correct for both measured and unmeasured pleiotropy. Stat Med 36, 4705–4718 (2017).

68 Burgess, S., Dudbridge, F. & Thompson, S. G. Re: “Multivariable Mendelian randomization: the use of pleiotropic genetic variants to estimate causal effects”. Am J Epidemiol 181, 290–291 (2015).

69 Yavorska, O. O. & Burgess, S. MendelianRandomization: an R package for performing Mendelian randomization analyses using summarized data. Int J Epidemiol 46, 1734–1739 (2017).

70 Mitchell, R., Elsworth, BL, Mitchell, R, Raistrick, CA, Paternoster, L, Hemani, G, Gaunt, TR. (2019).

