## Supplementary Information for "Investigating the effect of sexual behaviour on oropharyngeal cancer risk: a methodological assessment of Mendelian randomization"

**Methods**

*Mendelian randomization sensitivity analyses*

In addition to application of the inverse variance weighted (IVW) method (which combines Wald ratios for each SNP together in fixed effect meta-analysis) for Mendelian randomization (MR), weighted median ^1^, weighted mode ^2^ and MR-Egger ^3^ were also used. Whereas IVW assumes all SNP-exposure associations are known, the weighted median stipulates that at least 50% of the weight in the analysis stems from valid instruments ^1^, while the weighted mode requires the largest subset of instruments which identify the same effect to be valid instruments ^2^. MR-Egger can provide unbiased estimates even when all SNPs in an instrument affect the outcome by means other than via the risk factor of interest (i.e., violating the ‘exclusion restriction assumption’ of MR). However, MR-Egger assumes negligible measurement error (NOME) ^4^ in the genetic instrument and that the InSIDE (Instrument Strength Independent of Direct Effect) assumption remains satisfied ^3^.

Where there was evidence of violation of the no measurement error (NOME) assumption, assessed based on the I^2^ statistic (<0.90), MR-Egger was performed with simulation extrapolation (SIMEX) correction ^4^. To further assess the robustness of our findings, we inspected the Cochran’s Q statistic, which assesses heterogeneity between individual genetic variants, and which may indicate the presence of invalid instruments, for example due to horizontal pleiotropy. Scatter and leave-one-out plots were produced to evaluate influential outliers and Mendelian Randomization Pleiotropy RESidual Sum and Outlier (MR-PRESSO) was applied to detect and correct for potential outliers (*p* <0.05) ^5^.

*Genome-wide association study for HPV and C. trachomatis seropositivity*

A pilot of 9,695 participants in UK Biobank had serological measures using a multiplex serology panel (IgG) ^6^. UK Biobank is a large population-based cohort study that recruited over 500,000 men and women aged between 37 and 73 years between 2006 and 2010 throughout the UK ^7^. It received ethical approval from the National Health Service North West Centre for Research Ethics Committee (reference: 11/NW/0382). UK Biobank approval was given for this project (ID 40644) and UK Biobank GWAS data was also accessed under the application (ID 15825). Details of genotyping quality control, phasing and imputation are described elsewhere ^7^.

Multiplex serology was performed at serum dilution 1:1000 using previously developed methodology ^8,9^ and applied in various epidemiological studies, including studies on oropharyngeal cancer ^10,11^. The output from the Luminex reader produced quantitative data expressed in median fluorescence intensity (MFI) values per pathogen-specific antigen and serum. Seropositivity for each antigen was determined by examining percentile plots and cut-offs were chosen to optimise sensitivity and specificity, based on validation work ^12^.

Details of the variants and samples quality control procedures are described in Kachuri *et al.* ^13^. Briefly, GWAS was performed using PLINK 2.0 (July 27, 2020, version), restricting to individuals of predominantly European ancestry, based on self-report and genetic ancestry principal components, without any immunodeficiencies. For the present analysis we further excluded anyone diagnosed with cancers of the oral cavity (n=12) and oropharynx (n=6), leaving a total of 7,906 individuals. Four HPV seropositivity markers were derived from UK Biobank using the following MFI cut-offs: HPV16 L1 antigen >175; HPV16 E6 antigen >120; HPV16 E7 antigen >150; HPV18 L1 antigen >175. Seropositivity GWAS were performed for HPV18 L1 and HPV16 L1, E6, and E7 proteins separately. For C. trachomatis seropositivity we focused on the pGP3 antigen (MFI>200; 1,609 seropositive cases), which is considered to be the gold standard for detecting current or past chlamydia infections compared to other less persistent antigens ^14,15^. GWAS were run using logistic regression in ‘firth-fallback’ mode to implement Firth regression if the standard model failed to converge, particularly due to sparse cell counts. Models were adjusted for the top 10 genetic principal components, age at sample collection, genotyping array, sex, serology assay date, and quality control flag (indicating potential sample spill-over or an extra freeze/thaw cycle).

*Positive and negative control analyses*

Cervical cancer cases in the UK Biobank were identified using linked cancer registry records, as previously described in Graff *et al* ^16^. Analyses were restricted to malignant cervical cancers based on ICD codes for the type of cancer (ICD-10: C530, C531, C538, C539; ICD-9: 1800, 1801, 1808, 1809). We excluded diagnoses with carcinoma in situ, benign, or uncertain behaviour, however cases with missing behaviour codes were retained. Controls were restricted to individuals who had no record of cancer in the registry data, did not self-report a prior history of cancer (other than non-melanoma skin cancer), and, if deceased, who did not have cancer listed as a cause of death. Individuals with discordant self-reported and genetic sex and sex-discordant cancers were excluded. The analytic dataset was restricted to individuals of predominantly European ancestry, which was identified by self-report and refined by excluding samples with any of the first two genetic ancestry principal components (PCs) outside of 5 standard deviations (SD) of the population mean, leaving 955 cases and 187,257 controls. GWAS was performed using logistic regression with adjustment for age, genotyping array, and the top 10 PCs using PLINK 2.0 (July 27, 2020, version) in ‘firth-fallback’ mode.

For lung cancer, a GWAS of 11,348 cases and 15,861 controls which was conducted under the auspices of the Transdisciplinary Research In Cancer of the Lung (TRICL) Research Team, part of the GAME-ON consortium and associated with the International Lung Cancer Consortium (ILCCO) ^17^. Oral cancer cases (n=2,990) and 6,585 controls were taken from the Genetic Associations and Mechanisms in Oncology (GAME-ON) Network using ICD-10 codes: oral cavity (C02.0–C02.9, C03.0–C03.9, C04.0–C04.9 and C05.0–C06.9). Details of the studies included as well as the genotyping and imputation performed along with the GWAS conducted, have been described previously ^18,19^.

**Supplementary information**

**Supplementary Table 1.** Univariable Mendelian randomization results of age at first sex with HPV seropositivity including sensitivity analyses.

| **Outcome** | **Method** | **OR** | **CIL** | **CIU** | ***P*-value** |
| --- | --- | --- | --- | --- | --- |
| HPV16 L1 | Inverse variance weighted | 0.47 | 0.21 | 1.01 | 0.05 |
| HPV16 L1 | MR-Egger | 0.05 | 2.26E-03 | 1.18 | 0.07 |
| HPV16 L1 | Weighted median | 0.35 | 0.11 | 1.09 | 0.07 |
| HPV16 L1 | Weighted mode | 0.11 | 4.69E-03 | 2.76 | 0.18 |
| HPV16 E6 | Inverse variance weighted | 1.35 | 0.38 | 4.74 | 0.64 |
| HPV16 E6 | MR-Egger | 0.66 | 4.24E-03 | 1.02E+02 | 0.87 |
| HPV16 E6 | Weighted median | 0.51 | 0.08 | 3.27 | 0.48 |
| HPV16 E6 | Weighted mode | 0.02 | 9.38E-05 | 5.42 | 0.18 |
| HPV16 E7 | Inverse variance weighted | 1.51 | 0.62 | 3.66 | 0.36 |
| HPV16 E7 | MR-Egger | 0.16 | 4.66E-03 | 5.65 | 0.32 |
| HPV16 E7 | Weighted median | 1.40 | 0.37 | 5.31 | 0.62 |
| HPV16 E7 | Weighted mode | 1.24 | 0.02 | 67.5E | 0.92 |
| HPV18 L1 | Inverse variance weighted | 0.76 | 0.28 | 2.10 | 0.60 |
| HPV18 L1 | MR-Egger | 0.47 | 0.01 | 26.72 | 0.71 |
| HPV18 L1 | Weighted median | 0.79 | 0.18 | 3.58 | 0.76 |
| HPV18 L1 | Weighted mode | 0.34 | 0.01 | 22.65 | 0.62 |

Abbreviations: IVW, inverse variance weighted; SE, standard error; OR, odds ratio; CI, confidence intervals; SNPs, single nucleotide polymorphisms. OR represents the exponential change in odds of HPV seropositivity per SD change (7.3-month delay) in age at first sex. GWAS were run for four HPV markers derived from UK Biobank, with HPV16 seropositivity described: if antigen L1 >175; if antigen E6 >120 or antigen E7 >150.

**Supplementary Table 2.** Mendelian randomization results of number of sexual partners with HPV seropositivity including sensitivity analyses.

| **Outcome** | **Method** | **Beta** | **SE** | **OR** | **CIL** | **CIU** | ***P*-value** |
| --- | --- | --- | --- | --- | --- | --- | --- |
| HPV16 L1 | Inverse variance weighted | 0.73 | 0.53 | 2.07 | 0.74 | 5.83 | 0.17 |
| HPV16 L1 | MR-Egger | -0.56 | 2.37 | 0.57 | 0.01 | 58.80 | 0.81 |
| HPV16 L1 | Weighted median | 0.95 | 0.71 | 2.58 | 0.64 | 10.40 | 0.18 |
| HPV16 L1 | Weighted mode | 2.92 | 1.92 | 18.50 | 0.43 | 795.00 | 0.13 |
| HPV16 E6 | Inverse variance weighted | 1.68 | 0.85 | 5.39 | 1.03 | 28.30 | 0.05 |
| HPV16 E6 | MR-Egger | 3.01 | 3.79 | 20.40 | 0.01 | 3.42E+04 | 0.43 |
| HPV16 E6 | Weighted median | 1.59 | 1.21 | 4.91 | 0.46 | 52.50 | 0.19 |
| HPV16 E6 | Weighted mode | 5.45 | 3.57 | 2.33E+02 | 0.21 | 2.55E+05 | 0.13 |
| HPV16 E7 | Inverse variance weighted | -0.12 | 0.58 | 0.89 | 0.28 | 2.80 | 0.84 |
| HPV16 E7 | MR-Egger | -1.64 | 2.56 | 0.19 | <0.001 | 29.30 | 0.52 |
| HPV16 E7 | Weighted median | -0.43 | 0.86 | 0.65 | 0.12 | 3.49 | 0.61 |
| HPV16 E7 | Weighted mode | -0.73 | 2.45 | 0.48 | <0.001 | 58.50 | 0.76 |
| HPV18 L1 | Inverse variance weighted | -0.76 | 0.67 | 0.47 | 0.13 | 1.73 | 0.25 |
| HPV18 L1 | MR-Egger | -6.74 | 2.89 | <0.001 | <0.001 | 0.34 | 0.02 |
| HPV18 L1 | Weighted median | -0.24 | 0.98 | 0.78 | 0.11 | 5.40 | 0.80 |
| HPV18 L1 | Weighted mode | 1.73 | 2.67 | 5.67 | 0.03 | 1.07E+03 | 0.52 |

Abbreviations: IVW, inverse variance weighted; SE, standard error; OR, odds ratio; CI, confidence intervals; SNPs, single nucleotide polymorphisms. OR represents the exponential change in odds of HPV seropositivity per SD increase (0.94) in number of sexual partners. GWAS were run for four HPV markers derived from UK Biobank, with HPV16 seropositivity described: if antigen L1 >175; if antigen E6 >120 or antigen E7 >150.

**Supplementary Table 3.** Assessing weak instrument bias (F-statistic) and proportion of variance in the phenotype (*R*^2^) explained by age at first sex and number of sexual partners genetic instruments.

|  | **N SNPs** | **R^2^** | **F-statistic** |
| --- | --- | --- | --- |
| **AFS** | 139 | 0.020 | 47 |
| **NSP** | 117 | 0.012 | 40 |

Abbreviations: AFS, age at first sex; NSP, number of sexual partners.

**Supplementary Table 4.** Assessing heterogeneity of single nucleotide polymorphism effect estimates in inverse variance weighted and MR-Egger regression for univariable MR analysis of age at first sex and number of sexual partners on oropharyngeal cancer risk.

| **Exposure** | **Outcome** | **Exposure dataset** | **Q IVW** | **df** | ***P*** | **Q MR-Egger** | **df** | ***P*** |
| --- | --- | --- | --- | --- | --- | --- | --- | --- |
| **AFS** | Oropharyngeal cancer | UK Biobank^20^ | 159.42 | 138 | 0.102 | 158.64 | 137 | 0.099 |
| **NSP** | Oropharyngeal cancer | UK Biobank^21^ | 155.57 | 115 | 0.007 | 155.55 | 114 | 0.006 |

Abbreviations: AFS, age at first sex; NSP, number of sexual partners; Q, Cochran’s Q-statistic; df, degrees of freedom; P, p-value.

**Supplementary Table 5.** Assessing directional pleiotropy through MR-Egger intercept for univariable MR analysis of age at first sex and number of sexual partners on oropharyngeal cancer risk.

| **Exposure** | **Outcome** | **Exposure dataset** | **N SNPs** | **Estimate** | **SE** | **P** |
| --- | --- | --- | --- | --- | --- | --- |
| **AFS** | Oropharyngeal cancer | UK Biobank^20^ | 139 | 0.012 | 0.015 | 0.42 |
| **NSP** | Oropharyngeal cancer | UK Biobank^21^ | 117 | -0.003 | 0.021 | 0.89 |

Abbreviations: AFS, age at first sex; NSP, number of sexual partners; SE, standard error; P, p-value.

**Supplementary Table 6.** MR-PRESSO outliers detected results for age at first sex and number of sexual partners instruments on oropharyngeal cancer risk.

|  | **SNP** | **Q-stat** | ***P*-value** | **Q-sum** | **Q-difference** |
| --- | --- | --- | --- | --- | --- |
| **AFS** | rs28929474 | 7.64 | 0.01 | 7.64 | -151.43 |
|  | rs2406374 | 7.61 | 0.01 | 15.25 | -143.82 |
|  | rs783544 | 6.87 | 0.01 | 22.12 | -136.95 |
|  | rs1925686 | 5.27 | 0.02 | 27.39 | -131.68 |
|  | rs222440 | 4.75 | 0.03 | 32.14 | -126.93 |
|  | rs12701263 | 4.69 | 0.03 | 36.83 | -122.24 |
|  | rs4800204 | 4.38 | 0.04 | 41.21 | -117.86 |
|  | rs875097 | 4.08 | 0.04 | 45.28 | -113.79 |
| **NSP** | rs446952 | 10.17 | <0.001 | 10.17 | -145.15 |
|  | rs10138911 | 7.60 | 0.01 | 17.76 | -137.55 |
|  | rs357508 | 7.15 | 0.01 | 24.91 | -130.40 |
|  | rs35219418 | 6.38 | 0.01 | 31.29 | -124.02 |
|  | rs2422136 | 5.66 | 0.02 | 36.95 | -118.36 |
|  | rs34488670 | 4.66 | 0.03 | 41.61 | -113.71 |
|  | rs62063281 | 4.10 | 0.04 | 45.71 | -109.61 |

Abbreviations: AFS, age at first sex; NSP, number of sexual partners; Q-stat, Cochran’s Q statistic.

**Supplementary Table 7.** MR-PRESSO results for age at first sex and number of sexual partners instruments on oropharyngeal cancer risk.

| **Outcome** | **Exposure** | **RSSobs** | ***P*-value** |
| --- | --- | --- | --- |
| Oropharyngeal cancer | **AFS** | 199.47 | 0.07 |
| Oropharyngeal cancer | **NSP** | 158.15 | 0.01 |

Abbreviations: AFS, age at first sex; NSP, number of sexual partners; RSSobs, residual sum of squares observations.

**Supplementary Table 8.** Outlier corrected results for age at first sex and number of sexual partners instruments on combined oropharyngeal cancer.

| **Outcome** | **Exposure** | **N SNPs** | **Method** | **OR** | **CIL** | **CIU** | ***P*-value** |
| --- | --- | --- | --- | --- | --- | --- | --- |
| Oropharyngeal cancer | AFS | 131 | IVW | 0.37 | 0.24 | 0.58 | <0.001 |
| Oropharyngeal cancer | AFS | 131 | MR-Egger | 0.39 | 0.06 | 2.39 | 0.31 |
| Oropharyngeal cancer | AFS | 131 | Weighted median | 0.36 | 0.19 | 0.67 | <0.001 |
| Oropharyngeal cancer | AFS | 131 | Weighted mode | 0.21 | 0.03 | 1.37 | 0.11 |
| Oropharyngeal cancer | NSP | 109 | IVW | 2.08 | 1.34 | 3.25 | <0.001 |
| Oropharyngeal cancer | NSP | 109 | MR-Egger | 1.77 | 0.20 | 15.57 | 0.61 |
| Oropharyngeal cancer | NSP | 109 | Weighted median | 2.38 | 1.22 | 4.63 | 0.01 |
| Oropharyngeal cancer | NSP | 109 | Weighted mode | 3.18 | 0.48 | 21.33 | 0.24 |

Abbreviations: IVW, inverse variance weighted; SE, standard error; OR, odds ratio; CI, confidence intervals; SNPs, single nucleotide polymorphisms. NSP OR represents the exponential change in odds of oropharyngeal squamous cell carcinoma per SD increase (0.94) in number of sexual partners. AFS OR represents the exponential change in odds of oropharyngeal squamous cell carcinoma per SD change (7.3-month delay) in age at first sex.

**Supplementary Table 9.** Assessing violation of the “NO Measurement Error” (NOME) assumption for instruments used in MR-Egger regression of age at first sex and number of sexual partners on oropharyngeal cancer risk.

| **Exposure** | **Exposure dataset** | **I^2^ unweighted** |
| --- | --- | --- |
| **AFS** | UK Biobank^20^ | 0.61 |
| **NSP** | UK Biobank^21^ | 0.44 |

Abbreviations: I^2^, I-squared statistic.

**Supplementary Table 10.** SIMEX correction MR-Egger regression results for age at first sex and number of sexual partners instruments on oropharyngeal cancer risk (where I^2^ <0.90).

| **Outcome** | **Exposure** | **OR** | **CIL** | **CIU** | ***P*** |
| --- | --- | --- | --- | --- | --- |
| Oropharyngeal cancer | **AFS** | 0.01 | 0.001 | 0.14 | <0.001 |
| Oropharyngeal cancer | **NSP** | 3.63 | 0.41 | 32.11 | 0.25 |

Abbreviations: AFS, age at first sex; NSP, number of sexual partners; OR, odds ratio; CI, confidence intervals; *P*, p-value.

**Supplementary Table 11.** Univariable Mendelian randomization examining effects of age at first sex on positive and negative controls.

|  | **Outcome** | **Exposure** | **Method** | **OR** | **CIL** | **CIU** | **P** |
| --- | --- | --- | --- | --- | --- | --- | --- |
| Positive controls | Cervical cancer | AFS | Inverse variance weighted | 0.43 | 0.27 | 0.66 | <0.001 |
|  | Cervical cancer | AFS | MR-Egger | 0.45 | 0.07 | 2.67 | 0.38 |
|  | Cervical cancer | AFS | Weighted median | 0.52 | 0.27 | 1.00 | 0.05 |
|  | Cervical cancer | AFS | Weighted mode | 0.84 | 0.12 | 6.08 | 0.86 |
|  | C. trachomatis seropositivity | AFS | Inverse variance weighted | 0.40 | 0.27 | 0.60 | <0.001 |
|  | C. trachomatis seropositivity | AFS | MR-Egger | 0.10 | 0.02 | 0.50 | 0.01 |
|  | C. trachomatis seropositivity | AFS | Weighted median | 0.40 | 0.22 | 0.71 | <0.001 |
|  | C. trachomatis seropositivity | AFS | Weighted mode | 0.33 | 0.07 | 1.62 | 0.17 |
| Negative controls | Lung cancer | AFS | Inverse variance weighted | 0.12 | 0.05 | 0.29 | <0.001 |
|  | Lung cancer | AFS | MR-Egger | 0.83 | 0.02 | 27.47 | 0.91 |
|  | Lung cancer | AFS | Weighted median | 0.17 | 0.05 | 0.58 | <0.001 |
|  | Lung cancer | AFS | Weighted mode | 0.14 | 0.01 | 3.12 | 0.22 |
|  | Oral cancer | AFS | Inverse variance weighted | 0.63 | 0.41 | 0.96 | 0.03 |
|  | Oral cancer | AFS | MR-Egger | 0.56 | 0.10 | 3.25 | 0.52 |
|  | Oral cancer | AFS | Weighted median | 0.80 | 0.46 | 1.41 | 0.45 |
|  | Oral cancer | AFS | Weighted mode | 0.87 | 0.26 | 2.90 | 0.82 |

Abbreviations: SE, standard error; OR, odds ratio; P, *p*-value; CI, confidence intervals; AFS, age at first sex. AFS OR represents the exponential change in odds of cervical or lung cancer per SD change (7.3-month delay) in age at first sex.

**Supplementary Table 12.** Univariable Mendelian randomization examining effects of number of sexual partners on positive and negative controls.

|  | **Outcome** | **Exposure** | **Method** | **OR** | **CIL** | **CIU** | **P** |
| --- | --- | --- | --- | --- | --- | --- | --- |
| Positive controls | Cervical cancer | NSP | Inverse variance weighted | 1.94 | 0.97 | 3.91 | 0.06 |
|  | Cervical cancer | NSP | MR-Egger | 6.16 | 0.26 | 147.18 | 0.26 |
|  | Cervical cancer | NSP | Weighted median | 1.24 | 0.46 | 3.35 | 0.67 |
|  | Cervical cancer | NSP | Weighted mode | 0.58 | 0.05 | 6.81 | 0.67 |
|  | C. trachomatis seropositivity | NSP | Inverse variance weighted | 2.41 | 1.41 | 4.11 | <0.001 |
|  | C. trachomatis seropositivity | NSP | MR-Egger | 0.40 | 0.04 | 4.22 | 0.45 |
|  | C. trachomatis seropositivity | NSP | Weighted median | 1.97 | 0.91 | 4.28 | 0.09 |
|  | C. trachomatis seropositivity | NSP | Weighted mode | 2.97 | 0.36 | 24.74 | 0.32 |
| Negative controls | Lung cancer | NSP | Inverse variance weighted | 7.14 | 2.36 | 21.59 | <0.001 |
|  | Lung cancer | NSP | MR-Egger | 0.39 | <0.001 | 53.90 | 0.71 |
|  | Lung cancer | NSP | Weighted median | 10.26 | 2.63 | 40.04 | <0.001 |
|  | Lung cancer | NSP | Weighted mode | 18.82 | 0.61 | 578.05 | 0.10 |
|  | Oral cancer | NSP | Inverse variance weighted | 1.20 | 0.72 | 2.04 | 0.47 |
|  | Oral cancer | NSP | MR-Egger | 1.16 | 0.09 | 15.31 | 0.91 |
|  | Oral cancer | NSP | Weighted median | 1.52 | 0.81 | 2.84 | 0.19 |
|  | Oral cancer | NSP | Weighted mode | 1.96 | 0.38 | 10.01 | 0.42 |

Abbreviations: SE, standard error; OR, odds ratio; P, *p*-value; CI, confidence intervals; NSP, number of sexual partners; NSP OR represents the exponential change in odds of cervical or lung cancer per SD increase (0.94) in number of sexual partners.

**Supplementary Table 13.** Assessing directional pleiotropy through MR-Egger intercept for univariable MR positive and negative control analyses.

| **Exposure** | **Outcome** | **Exposure dataset** | **Estimate** | **SE** | **P** |
| --- | --- | --- | --- | --- | --- |
| **AFS** | Cervical cancer | UK Biobank | -0.001 | 0.014 | 0.96 |
| **NSP** | Cervical cancer | UK Biobank | -0.020 | 0.028 | 0.47 |
| **AFS** | C. trachomatis seropositivity | UK Biobank | 0.022 | 0.013 | 0.08 |
| **NSP** | C. trachomatis seropositivity | UK Biobank | 0.031 | 0.021 | 0.13 |
| **AFS** | Lung cancer | UK Biobank | -0.031 | 0.028 | 0.28 |
| **NSP** | Lung cancer | UK Biobank | 0.051 | 0.042 | 0.24 |
| **AFS** | Oral cancer | GAME-ON | 0.002 | 0.014 | 0.90 |
| **NSP** | Oral cancer | GAME-ON | 0.0007 | 0.022 | 0.97 |

Abbreviations: AFS, age at first sex; NSP, number of sexual partners; SE, standard error; P, p-value.

**Supplementary Table 14.** Assessing heterogeneity of single nucleotide polymorphism effect estimates in inverse variance weighted and MR-Egger regression for univariable MR positive and negative control analyses.

| **Exposure** | **Outcome** | **Exposure dataset** | **Q IVW** | **df** | ***P*** | **Q MR-Egger** | **df** | ***P*** |
| --- | --- | --- | --- | --- | --- | --- | --- | --- |
| **AFS** | Cervical cancer | UK Biobank | 211.73 | 219 | 0.625 | 211.73 | 218 | 0.607 |
| **NSP** | Cervical cancer | UK Biobank | 105.10 | 88 | 0.103 | 104.46 | 87 | 0.098 |
| **AFS** | C. trachomatis seropositivity | UK Biobank | 199.82 | 215 | 0.764 | 196.79 | 214 | 0.795 |
| **NSP** | C. trachomatis seropositivity | UK Biobank | 120.06 | 109 | 0.221 | 117.51 | 108 | 0.250 |
| **AFS** | Lung cancer | UK Biobank | 209.28 | 179 | 0.060 | 207.88 | 178 | 0.062 |
| **NSP** | Lung cancer | UK Biobank | 148.86 | 108 | 0.006 | 146.92 | 107 | 0.006 |
| **AFS** | Oral cancer | GAME-ON | 161.92 | 138 | 0.080 | 161.90 | 137 | 0.072 |
| **NSP** | Oral cancer | GAME-ON | 123.42 | 90 | 0.011 | 123.42 | 89 | 0.009 |

Abbreviations: AFS, age at first sex; NSP, number of sexual partners; Q, Cochran’s Q-statistic; IVW, inverse variance weighted; df, degrees of freedom; P, p-value.

**Supplementary Table 15.** MR-PRESSO outliers detected results for age at first sex and number of sexual partners instruments on positive and negative controls.

|  | **SNP** | **Q-stat** | **P-value** | **Q-sum** | **Q-difference** |
| --- | --- | --- | --- | --- | --- |
| **Lung cancer outliers** | | | | | |
| **AFS** | rs2910032 | 10.08 | <0.01 | 10.08 | -198.50 |
|  | rs6764919 | 9.09 | <0.01 | 19.17 | -189.42 |
|  | rs1156981 | 7.79 | 0.01 | 26.96 | -181.63 |
|  | rs147725178 | 7.29 | 0.01 | 34.24 | -174.34 |
|  | rs4804512 | 7.27 | 0.01 | 41.51 | -167.07 |
|  | rs9538248 | 5.62 | 0.02 | 47.14 | -161.45 |
|  | rs13280592 | 4.97 | 0.03 | 52.11 | -156.48 |
|  | rs57945129 | 4.86 | 0.03 | 56.97 | -151.61 |
|  | rs10134692 | 4.84 | 0.03 | 61.82 | -146.77 |
|  | rs2406374 | 4.58 | 0.03 | 66.40 | -142.19 |
|  | rs1368546 | 4.52 | 0.03 | 70.92 | -137.67 |
|  | rs7955865 | 4.45 | 0.03 | 75.37 | -133.21 |
|  | rs11204771 | 4.36 | 0.04 | 79.73 | -128.86 |
|  | rs9886840 | 4.25 | 0.04 | 83.98 | -124.60 |
|  | rs2274568 | 4.18 | 0.04 | 88.17 | -120.42 |
|  | rs198310 | 3.95 | 0.05 | 92.11 | -116.48 |
| **NSP** | rs34488670 | 9.15 | 0.01 | 9.15 | -139.13 |
|  | rs9570035 | 7.71 | 0.01 | 16.86 | -131.42 |
|  | rs2021638 | 6.61 | 0.01 | 23.47 | -124.81 |
|  | rs62519841 | 6.47 | 0.01 | 29.94 | -118.34 |
|  | rs6916079 | 6.24 | 0.01 | 36.18 | -112.10 |
|  | rs266060 | 5.63 | 0.02 | 41.80 | -106.47 |
|  | rs16918024 | 5.20 | 0.02 | 47.00 | -101.27 |
|  | rs716936 | 4.81 | 0.03 | 51.81 | -96.47 |
|  | rs9856718 | 4.74 | 0.03 | 56.56 | -91.72 |
|  | rs72780746 | 4.56 | 0.03 | 61.12 | -87.16 |
|  | rs2568464 | 4.51 | 0.03 | 65.63 | -82.65 |
|  | rs1145247 | 4.34 | 0.04 | 69.96 | -78.32 |
| **Cervical cancer outliers** | | | | | |
| **AFS** | 2:45184405 | 6.09 | 0.01 | 6.09 | -205.33 |
|  | rs1925686 | 5.80 | 0.02 | 11.89 | -199.54 |
|  | rs28406364 | 5.71 | 0.02 | 17.60 | -193.82 |
|  | rs56066200 | 5.52 | 0.02 | 23.12 | -188.30 |
|  | rs1260580524 | 5.38 | 0.02 | 28.50 | -182.92 |
|  | rs138850767 | 5.36 | 0.02 | 33.86 | -177.56 |
|  | rs1585634 | 5.16 | 0.02 | 39.02 | -172.40 |
|  | rs767943 | 4.75 | 0.03 | 43.78 | -167.65 |
|  | rs1568188906 | 4.43 | 0.04 | 48.21 | -163.21 |
|  | rs7533341 | 4.26 | 0.04 | 52.47 | -158.96 |
|  | 2:105942069 | 4.13 | 0.04 | 56.60 | -154.83 |
|  | rs1417473851 | 4.06 | 0.04 | 60.66 | -150.76 |
|  | rs34155040 | 3.90 | 0.05 | 64.57 | -146.86 |
|  | rs73581580 | 3.89 | 0.05 | 68.46 | -142.97 |
| **NSP** | rs455425 | 7.96 | <0.001 | 7.96 | -97.02 |
|  | rs6467145 | 6.29 | 0.01 | 14.25 | -90.73 |
|  | rs58184964 | 5.58 | 0.02 | 19.83 | -85.15 |
|  | rs163327 | 4.92 | 0.03 | 24.75 | -80.23 |
| **Oral cancer** | | | | | |
| **AFS** | rs12203592 | 11.29 | <0.001 | 11.29 | -150.50 |
|  | rs7236339 | 10.18 | <0.001 | 21.47 | -140.32 |
|  | rs794375 | 5.75 | 0.02 | 27.22 | -134.57 |
|  | rs141547796 | 4.87 | 0.03 | 32.09 | -129.70 |
|  | rs147725178 | 4.73 | 0.03 | 36.81 | -124.98 |
|  | rs2406374 | 4.69 | 0.03 | 41.51 | -120.28 |
|  | rs2397678 | 4.66 | 0.03 | 46.16 | -115.63 |
|  | rs56392241 | 4.57 | 0.03 | 50.73 | -111.06 |
|  | rs1866710 | 4.11 | 0.04 | 54.84 | -106.95 |
|  | rs11688027 | 3.97 | 0.05 | 58.81 | -102.98 |
|  | rs34811474 | 3.96 | 0.05 | 62.77 | -99.02 |
| **NSP** | rs446952 | 12.98 | <0.001 | 12.98 | -110.42 |
|  | rs11664298 | 11.48 | <0.001 | 24.46 | -98.94 |
|  | rs40465 | 8.61 | <0.001 | 33.07 | -90.33 |
|  | rs6576006 | 5.21 | 0.02 | 38.28 | -85.12 |
|  | rs72780746 | 5.08 | 0.02 | 43.36 | -80.04 |
| **C. trachomatis seropositivity** | | | | | |
| **AFS** | 15:47677875_at_a | 8.09 | <0.001 | <0.001 | -191.30 |
|  | rs12147463 | 7.02 | 0.01 | 15.11 | -184.28 |
|  | rs2279574 | 5.83 | 0.02 | 20.93 | -178.46 |
|  | rs6504551 | 4.64 | 0.03 | 25.57 | -173.81 |
|  | 2:44108016_tctc_t | 4.44 | 0.04 | 30.02 | -169.37 |
|  | rs141547796 | 4.21 | 0.04 | 34.22 | -165.17 |
|  | 9:14522303_aattg_a | 4.14 | 0.04 | 38.36 | -161.03 |
|  | rs993700 | 4.00 | 0.05 | 42.36 | -157.03 |
|  | rs551086366 | 3.95 | 0.05 | 46.31 | -153.08 |
|  | rs807478 | 3.93 | 0.05 | 50.25 | -149.14 |
|  | rs2091377 | 3.90 | 0.05 | 54.14 | -145.24 |
| **NSP** | rs34488670 | 9.21 | <0.001 | 9.21 | -110.52 |
|  | rs72930774 | 8.93 | <0.001 | 18.15 | -101.59 |
|  | rs40465 | 8.79 | <0.001 | 26.93 | -92.80 |
|  | rs6452792 | 5.58 | 0.02 | 32.52 | -87.22 |
|  | rs2279829 | 5.29 | 0.02 | 37.81 | -81.93 |
|  | rs1106095 | 4.48 | 0.03 | 42.29 | -77.45 |
|  | rs75120545 | 4.37 | 0.04 | 46.66 | -73.07 |

Abbreviations: AFS, age at first sex; NSP, number of sexual partners; Q-stat, Cochran’s Q statistic.

**Supplementary Table 16.** MR-PRESSO results for age at first sex and number of sexual partners instruments on positive and negative controls.

| **Outcome** | **Exposure** | **RSSobs** | ***P*-value** |
| --- | --- | --- | --- |
| Cervical cancer | **AFS** | 213.63 | 0.63 |
| Cervical cancer | **NSP** | 107.54 | 0.10 |
| C. trachomatis seropositivity | **AFS** | 201.71 | 0.78 |
| C. trachomatis seropositivity | **NSP** | 122.56 | 0.22 |
| Lung cancer | **AFS** | 211.49 | 0.07 |
| Lung cancer | **NSP** | 151.49 | 0.01 |
| Oral cancer | **AFS** | 164.02 | 0.10 |
| Oral cancer | **NSP** | 126.08 | 0.01 |

Abbreviations: AFS, age at first sex; NSP, number of sexual partners; RSSobs, residual sum of squares observations.

**Supplementary Table 17.** Outlier corrected results for age at first sex and number of sexual partners instruments on lung and cervical cancer.

| **Outcome** | **Exposure** | **Method** | **OR** | **CIL** | **CIU** | ***P*-value** |
| --- | --- | --- | --- | --- | --- | --- |
| Cervical cancer | AFS | IVW | 0.42 | 0.27 | 0.67 | <0.001 |
| Cervical cancer | AFS | MR-Egger | 0.42 | 0.07 | 2.56 | 0.35 |
| Cervical cancer | AFS | Weighted median | 0.52 | 0.28 | 0.97 | 0.04 |
| Cervical cancer | AFS | Weighted mode | 0.86 | 0.14 | 5.46 | 0.88 |
| Cervical cancer | NSP | IVW | 1.91 | 1.00 | 3.68 | 0.05 |
| Cervical cancer | NSP | MR-Egger | 2.74 | 0.15 | 51.03 | 0.50 |
| Cervical cancer | NSP | Weighted median | 1.24 | 0.47 | 3.25 | 0.66 |
| Cervical cancer | NSP | Weighted mode | 0.61 | 0.05 | 7.81 | 0.70 |
| C. trachomatis seropositivity | AFS | IVW | 0.43 | 0.29 | 0.65 | <0.001 |
| C. trachomatis seropositivity | AFS | MR Egger | 0.35 | 0.06 | 1.92 | 0.23 |
| C. trachomatis seropositivity | AFS | Weighted median | 0.42 | 0.24 | 0.74 | <0.001 |
| C. trachomatis seropositivity | AFS | Weighted mode | 0.29 | 0.06 | 1.43 | 0.13 |
| C. trachomatis seropositivity | NSP | IVW | 2.02 | 1.19 | 3.43 | 0.01 |
| C. trachomatis seropositivity | NSP | MR Egger | 0.47 | 0.04 | 5.87 | 0.56 |
| C. trachomatis seropositivity | NSP | Weighted median | 1.85 | 0.88 | 3.92 | 0.11 |
| C. trachomatis seropositivity | NSP | Weighted mode | 3.80 | 0.43 | 33.57 | 0.23 |
| Lung cancer | AFS | IVW | 0.14 | 0.06 | 0.32 | <0.01 |
| Lung cancer | AFS | MR-Egger | 1.11 | 0.04 | 30.84 | 0.95 |
| Lung cancer | AFS | Weighted median | 0.18 | 0.06 | 0.59 | <0.01 |
| Lung cancer | AFS | Weighted mode | 0.15 | 0.01 | 2.85 | 0.21 |
| Lung cancer | NSP | IVW | 9.72 | 3.60 | 26.26 | <0.01 |
| Lung cancer | NSP | MR-Egger | 11.04 | 0.13 | 975.51 | 0.30 |
| Lung cancer | NSP | Weighted median | 12.88 | 3.09 | 53.71 | <0.01 |
| Lung cancer | NSP | Weighted mode | 17.94 | 0.58 | 557.97 | 0.10 |
| Oral cancer | AFS | IVW | 0.63 | 0.42 | 0.94 | 0.02 |
| Oral cancer | AFS | MR-Egger | 0.58 | 0.11 | 3.13 | 0.52 |
| Oral cancer | AFS | Weighted median | 0.80 | 0.45 | 1.43 | 0.45 |
| Oral cancer | AFS | Weighted mode | 0.88 | 0.24 | 3.24 | 0.85 |
| Oral cancer | NSP | IVW | 1.36 | 0.86 | 2.15 | 0.19 |
| Oral cancer | NSP | MR-Egger | 2.23 | 0.24 | 20.37 | 0.48 |
| Oral cancer | NSP | Weighted median | 1.54 | 0.78 | 3.04 | 0.21 |
| Oral cancer | NSP | Weighted mode | 1.95 | 0.39 | 9.70 | 0.41 |

Abbreviations: SE, standard error; OR, odds ratio; P, *p*-value; CI, confidence intervals; AFS, age at first sex; NSP, number of sexual partners. AFS OR represents the exponential change in odds of cervical or lung cancer per SD change (7.3-month delay) in age at first sex. NSP OR represents the exponential change in odds cervical or lung cancer per SD increase (0.94) in number of sexual partners.

**Supplementary Table 18.** Causal Analysis Using Summary Effect estimates (CAUSE) results for age at first sex on risk of oropharyngeal cancer.

| **Exposure** | **Outcome** | **Model 1** | **Model 2** | **ELPD** | **se** | ***γ*** | **η** | **q** | **q CIL** | **q CIU** | ***P*-value** |
| --- | --- | --- | --- | --- | --- | --- | --- | --- | --- | --- | --- |
| AFS | OPC | Null | Sharing | -6.90 | 3.50 | NA | -2.75 | 0.22 | 0.07 | 0.42 | 0.02 |
| AFS | OPC | Null | Causal | -7.20 | 4.40 | -0.85 | 0.62 | 0.07 | <0.001 | 0.31 | 0.05 |
| AFS | OPC | Sharing | Causal | -0.26 | 1.60 | NA | NA | NA | NA | NA | 0.44 |

Abbreviations: OPC, oropharyngeal cancer; ELPD, expected log pointwise posterior density; se, standard error; *γ* (gamma), estimate of causal effect if causal model is correct; η (eta), estimate of correlated pleiotropy; q, proportion of effect due to correlated pleiotropy; CI, confidence intervals; NA, non-applicable.

**Supplementary Table 19.** Causal Analysis Using Summary Effect estimates (CAUSE) results for number of sexual partners on risk of oropharyngeal cancer.

| **Exposure** | **Outcome** | **Model 1** | **Model 2** | **ELPD** | **se** | ***γ*** | **η** | **q** | **q CIL** | **q CIU** | ***P*-value** |
| --- | --- | --- | --- | --- | --- | --- | --- | --- | --- | --- | --- |
| NSP | OPC | Null | Sharing | -0.26 | 0.49 | NA | 1.79 | 0.08 | <0.001 | 0.34 | 0.30 |
| NSP | OPC | Null | Causal | -1.30 | 2.10 | 0.83 | -0.14 | 0.07 | <0.001 | 0.28 | 0.27 |
| NSP | OPC | Sharing | Causal | -1.10 | 1.70 | NA | NA | NA | NA | NA | 0.26 |

Abbreviations: OPC, oropharyngeal cancer; ELPD, expected log pointwise posterior density; se, standard error; *γ* (gamma), estimate of causal effect if causal model is correct; η (eta), estimate of correlated pleiotropy; q, proportion of effect due to correlated pleiotropy; CI, confidence intervals; NA, non-applicable

**Supplementary Table 20.** Overlapping single nucleotide polymorphisms identified between genetic instruments used in multivariable Mendelian randomization.

| **SNP** | **Genetic instrument** |
| --- | --- |
| rs10922907 | AFS->CSI |
| rs359243 | AFS->CSI |
| rs12244388 | AFS->CSI |
| rs3896224 | AFS->CSI |
| rs4702 | AFS->NSP |
| rs148544378 | AFS->NSP |
| rs6748341 | AFS->RT |
| rs7783012 | AFS->RT |
| rs28929474 | AFS->DPW |
| rs12517438 | AFS->SI |
| rs10853981 | AFS->SI |
| rs2279829 | NSP->SI |
| rs72780746 | NSP->SI |
| rs1531518 | NSP->RT |
| rs58400863 | RT->SI |
| rs6874731 | RT->SI |
| rs11783093 | CSI->SI |
| rs76608582 | CSI->SI |
| rs3811038 | CSI->SI |
| rs11768481 | CSI->SI |
| rs1050847 | CSI->SI |

Abbreviations: AFS, age at first sex; NSP, number of sexual partners; RT, risk tolerance; CSI, comprehensive smoking index; SI, smoking initiation; DPW, drinks per week.

**Supplementary Table 21.** LD Score Regression results for all exposures

| **Exposure 1** | **Exposure 2** | **rg** | **SE** | **z-score** | ***p*-value** | **h^2^ obs** | **h^2^ obs se** | **h^2^ int** | **h^2^ int se** | **gcov int** | **gcov int se** |
| --- | --- | --- | --- | --- | --- | --- | --- | --- | --- | --- | --- |
| AFS | NSP | -0.5946 | 0.014 | -42.4034 | 0 | 0.0993 | 0.0035 | 1.0088 | 0.0097 | -0.3328 | 0.009 |
| AFS | SI | -0.6169 | 0.0132 | -46.8167 | 0 | 0.0695 | 0.0023 | 0.8852 | 0.0095 | 0.1523 | 0.0092 |
| AFS | DPW | -0.1977 | 0.0182 | -10.8544 | 1.90E-27 | 0.0493 | 0.0022 | 0.9216 | 0.009 | 0.0717 | 0.0068 |
| AFS | RT | -0.4103 | 0.0222 | -18.5078 | 1.79E-76 | 0.0518 | 0.0024 | 1.0075 | 0.0097 | -0.0965 | 0.0087 |
| NSP | SI | 0.5298 | 0.0165 | 32.1864 | 2.74E-227 | 0.0693 | 0.0023 | 0.8858 | 0.0097 | -0.1319 | 0.0079 |
| NSP | DPW | 0.3494 | 0.0221 | 15.7754 | 4.59E-56 | 0.0491 | 0.0022 | 0.9221 | 0.009 | -0.1057 | 0.0069 |
| NSP | RT | 0.5912 | 0.0206 | 28.6502 | 1.59E-180 | 0.0519 | 0.0023 | 1.0059 | 0.0094 | 0.1727 | 0.0075 |
| SI | DPW | 0.4075 | 0.02 | 20.4162 | 1.20E-92 | 0.0492 | 0.0021 | 0.9224 | 0.0086 | 0.106 | 0.0061 |
| SI | RT | 0.327 | 0.0229 | 14.2622 | 3.76E-46 | 0.0517 | 0.0022 | 1.006 | 0.0093 | -0.0525 | 0.0074 |
| DPW | RT | 0.2859 | 0.0249 | 11.4728 | 1.81E-30 | 0.052 | 0.0023 | 1.0058 | 0.0094 | -0.0503 | 0.0063 |

Abbreviations: AFS, age at first sex; NSP, number of sexual partners; CSI, comprehensive smoking index; SI, smoking initiation; DPW, drinks per week; RT, risk tolerance; rg, genetic correlation; SE, bootstrap standard error of genetic correlation, h^2^ obs = estimated SNP heritability of the second exposure , h^2^ obs se = bootstrap standard error of the SNP heritability estimate, h^2^ int = LD score regression intercept for the second exposure, h^2^ int se = bootstrap standard error of the intercept, gcov int = estimated genetic covariance between exposure 1 and 2, gcov int se = bootstrap standard error of the genetic covariance

**Supplementary Table 22.** Assessing directional pleiotropy through MR-Egger intercept for multivariable MR analysis on oropharyngeal cancer.

| **Exposure** | **Outcome** | **Covariate** | **N SNPs** | **Estimate** | **SE** | **P** |
| --- | --- | --- | --- | --- | --- | --- |
| AFS | Oropharyngeal cancer | NSP | 152 | -1.44 | 0.01 | 0.35 |
| AFS | Oropharyngeal cancer | CSI | 174 | -0.34 | 0.01 | 0.40 |
| AFS | Oropharyngeal cancer | SI | 215 | -0.50 | 0.01 | 0.37 |
| AFS | Oropharyngeal cancer | DPW | 147 | -0.84 | 0.01 | 0.30 |
| AFS | Oropharyngeal cancer | RT | 160 | -1.41 | 0.01 | 0.10 |
| NSP | Oropharyngeal cancer | AFS | 152 | -1.44 | 0.01 | 0.35 |
| NSP | Oropharyngeal cancer | CSI | 157 | 1.70 | 0.01 | 0.81 |
| NSP | Oropharyngeal cancer | SI | 195 | 0.51 | 0.01 | 0.77 |
| NSP | Oropharyngeal cancer | DPW | 117 | 2.17 | 0.01 | 0.55 |
| NSP | Oropharyngeal cancer | RT | 125 | 0.75 | 0.01 | 0.92 |

Abbreviations: AFS, age at first sex; NSP, number of sexual partners; SE, standard error; P, p-value; CSI, comprehensive smoking index; SI, smoking initiation; DPW, drinks per week; RT, risk tolerance.

**Supplementary Table 23.** Multivariable Mendelian randomization for age at first sex and number of sexual partners with risk lung cancer.

| **Exposure** | **Exposure dataset** | **N SNPs** | **F-stat** | **Q-stat** | **P-value for instrument validity** | **Method** | **OR** | **95% CI** | ***P*** |
| --- | --- | --- | --- | --- | --- | --- | --- | --- | --- |
| **Age at first sex** | | | | | | | | | |
| Comprehensive Smoking Index | UK Biobank^22^ | 172 | 8.74 | 243.85 | <0.001 | IVW | 1.99 | 1.31, 3.02 | <0.001 |
|  | | | | | | MR-Egger | 1.15 | 0.58, 2.27 | 0.69 |
| Smoking initiation | GSCAN^23^ | 213 | 6.42 | 251.87 | 0.025 | IVW | 1.11 | 0.78, 1.58 | 0.57 |
|  | | | | | | MR-Egger | 1.34 | 0.77, 2.33 | 0.30 |
| Drinks per week | GSCAN^23^ | 145 | 29.38 | 162.58 | 0.11 | IVW | 0.74 | 0.57, 0.96 | 0.03 |
|  | | | | | | MR-Egger | 1.09 | 0.62, 1.93 | 0.76 |
| Risk tolerance | UK Biobank^21^ | 155 | 13.43 | 185.02 | 0.04 | IVW | 0.68 | 0.50, 0.92 | 0.01 |
|  | | | | | | MR-Egger | 0.71 | 0.39, 1.30 | 0.27 |
| **Number of sexual partners** | | | | | | | | | |
| Comprehensive Smoking Index | UK Biobank^22^ | 151 | 12.41 | 267.57 | <0.001 | IVW | 0.69 | 0.44, 1.07 | 0.09 |
|  | | | | | | MR-Egger | 0.79 | 0.46, 1.35 | 0.39 |
| Smoking initiation | GSCAN^23^ | 191 | 7.85 | 224.67 | 0.035 | IVW | 0.90 | 0.62, 1.31 | 0.58 |
|  | | | | | | MR-Egger | 1.46 | 0.88, 2.41 | 0.14 |
| Drinks per week | GSCAN^23^ | 112 | 22.62 | 144.72 | 0.013 | IVW | 1.61 | 1.14, 2.26 | 0.006 |
|  | | | | | | MR-Egger | 1.63 | 1.11, 2.40 | 0.013 |
| Risk tolerance | UK Biobank^21^ | 122 | 6.88 | 154.20 | 0.017 | IVW | 1.99 | 1.30, 3.05 | 0.002 |
|  | | | | | | MR-Egger | 2.40 | 1.28, 4.51 | 0.007 |

Abbreviations: IVW, inverse variance weighted; OR, odds ratio; CI, confidence intervals; *P, p*-value; Q-stat, Cochran’s Q statistic; F-stat, conditional F-statistic. AFS OR represents the exponential change in odds of oropharyngeal squamous cell carcinoma per SD change (7.3-month delay) in age at first sex. NSP OR represents the exponential change in odds of oropharyngeal squamous cell carcinoma per SD increase (0.94) in number of sexual partners.

**Supplementary Table 24.** Assessing directional pleiotropy through MR-Egger intercept for multivariable MR analysis on lung and cervical cancer.

| **Exposure** | **Outcome** | **Covariate** | **Estimate** | **SE** | **P** |
| --- | --- | --- | --- | --- | --- |
| AFS | Lung cancer | CSI | 0.011 | 0.005 | 0.05 |
| AFS | Lung cancer | SI | -0.003 | 0.004 | 0.38 |
| AFS | Lung cancer | DPW | -0.006 | 0.004 | 0.13 |
| AFS | Lung cancer | RT | -0.001 | 0.004 | 0.87 |
| NSP | Lung cancer | CSI | -0.006 | 0.006 | 0.34 |
| NSP | Lung cancer | SI | -0.008 | 0.003 | 0.01 |
| NSP | Lung cancer | DPW | 0.000 | 0.003 | 0.89 |
| NSP | Lung cancer | RT | -0.003 | 0.004 | 0.99 |

Abbreviations: AFS, age at first sex; NSP, number of sexual partners; SE, standard error; P, *p*-value; CSI, comprehensive smoking index; SI, smoking initiation; DPW, drinks per week; RT, risk tolerance.

**Supplementary Fig.1** Forest plots showing Mendelian randomization results for age at first sex and number of sexual partners single nucleotide polymorphisms with risk of oropharyngeal cancer in GAME-ON.

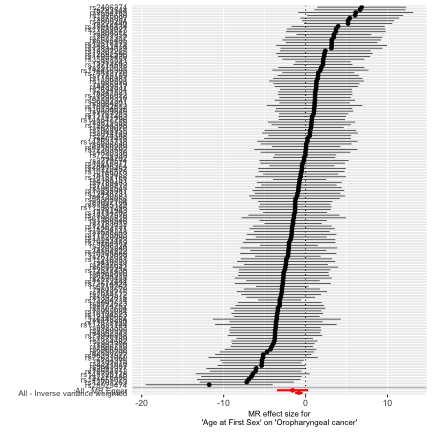
**A**

**B**

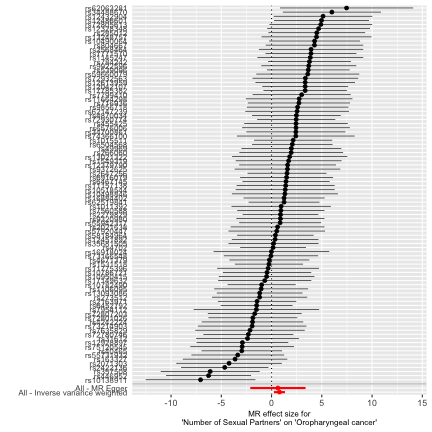

Effect estimates are reported on the log odds scale with 95% confidence intervals. **A** Age at first sex point estimate represents the exponential change in odds oropharyngeal squamous cell carcinoma per SD change (7.3 month delay) in age at first sex. **B** Number of sexual partners point estimate represents the exponential change in odds of oropharyngeal squamous cell carcinoma per SD increase (0.94) in number of sexual partners.

**Supplementary Fig.2** Scatter plots for age at first sex and number of sexual partners single nucleotide polymorphisms effect on oropharyngeal cancer in GAME-ON.

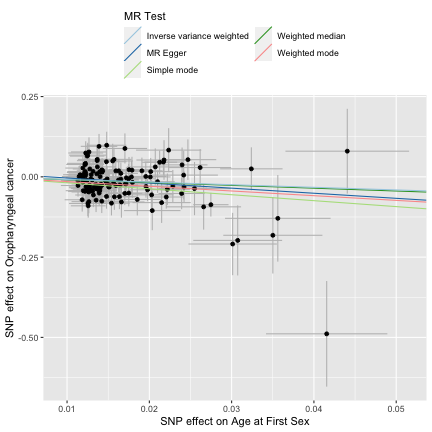

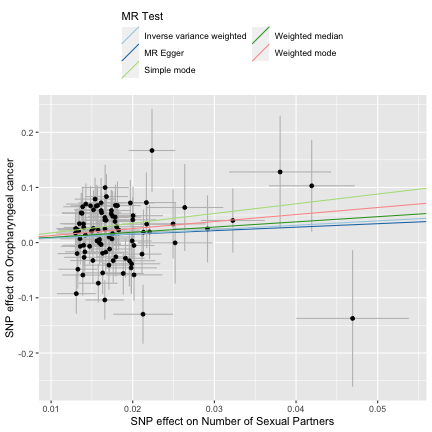

**Supplementary Fig.3** Leave one out plots for age at first sex and number of sexual partners single nucleotide polymorphisms effect on oropharyngeal cancer in GAME-ON.

**A**

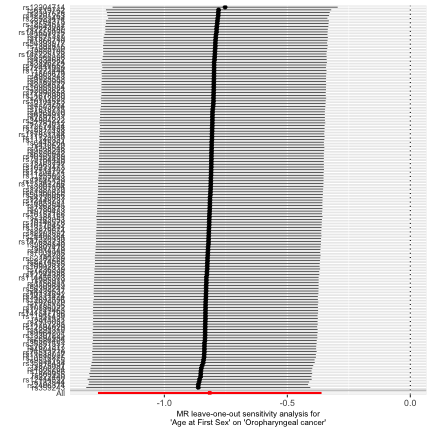

**B**
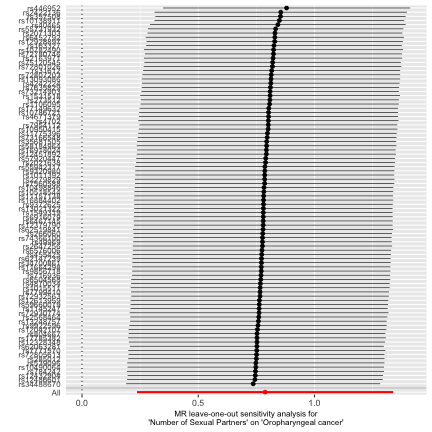

Leave one out plots for **A** age at first sex and **B** number of sexual partners.

**Supplementary Fig.4** Scatter and leave one out plots for age at first sex and number of sexual partners single nucleotide polymorphisms effect on risk of cervical cancer

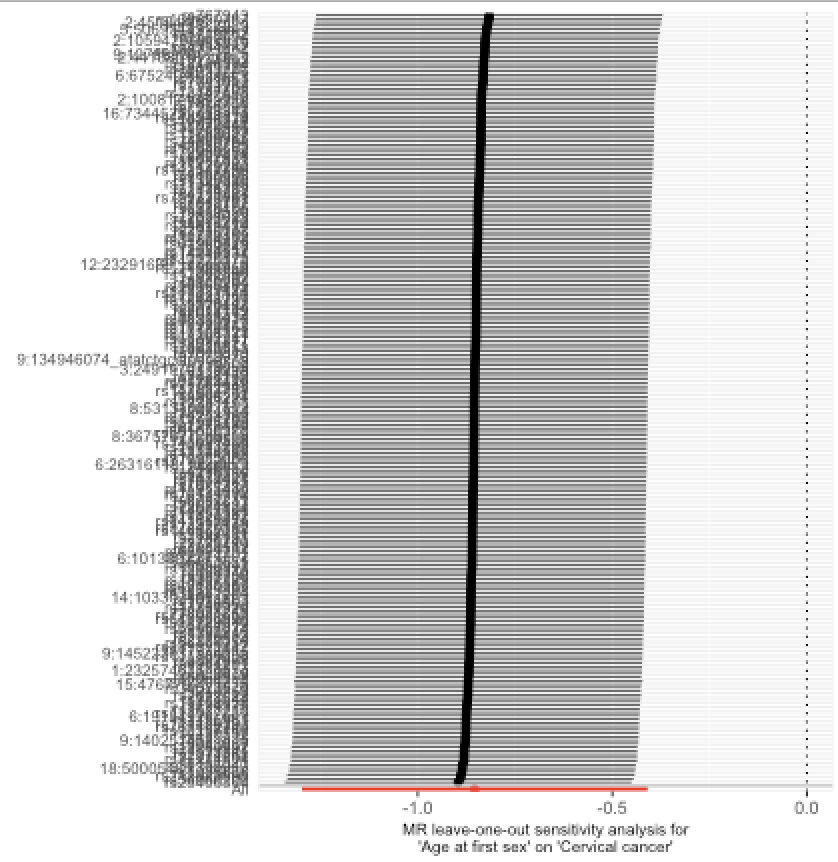

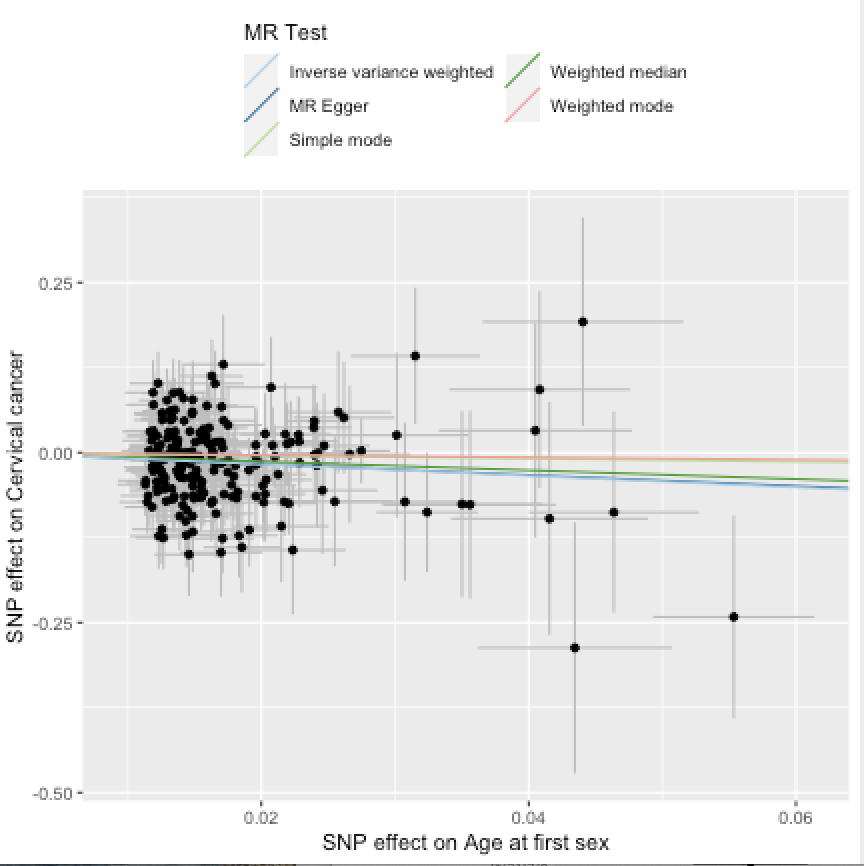

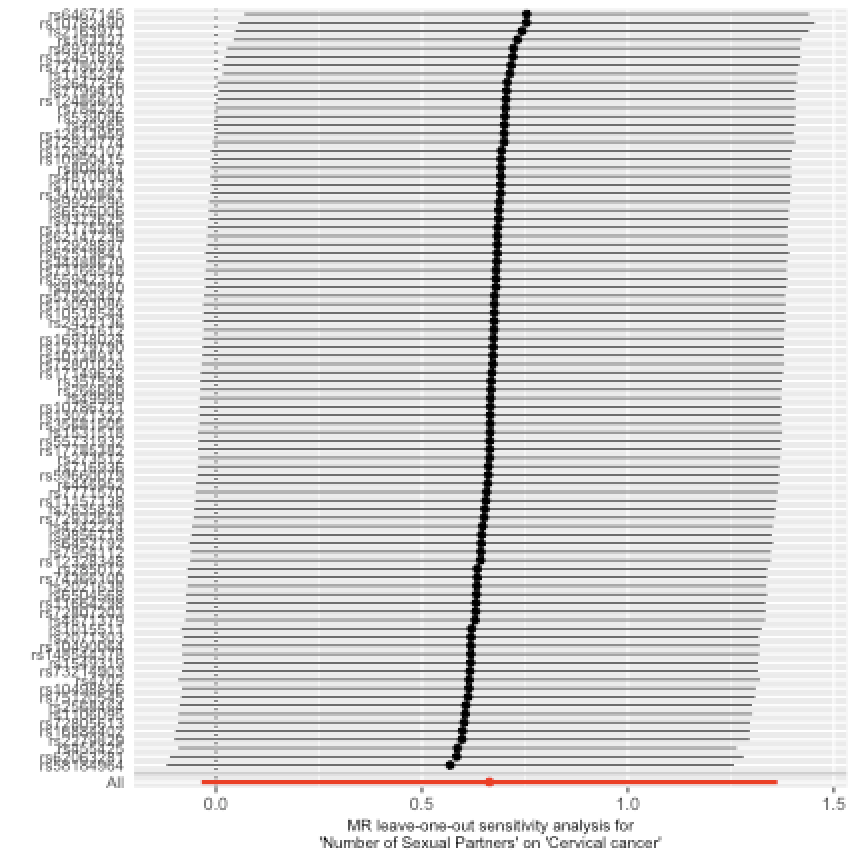

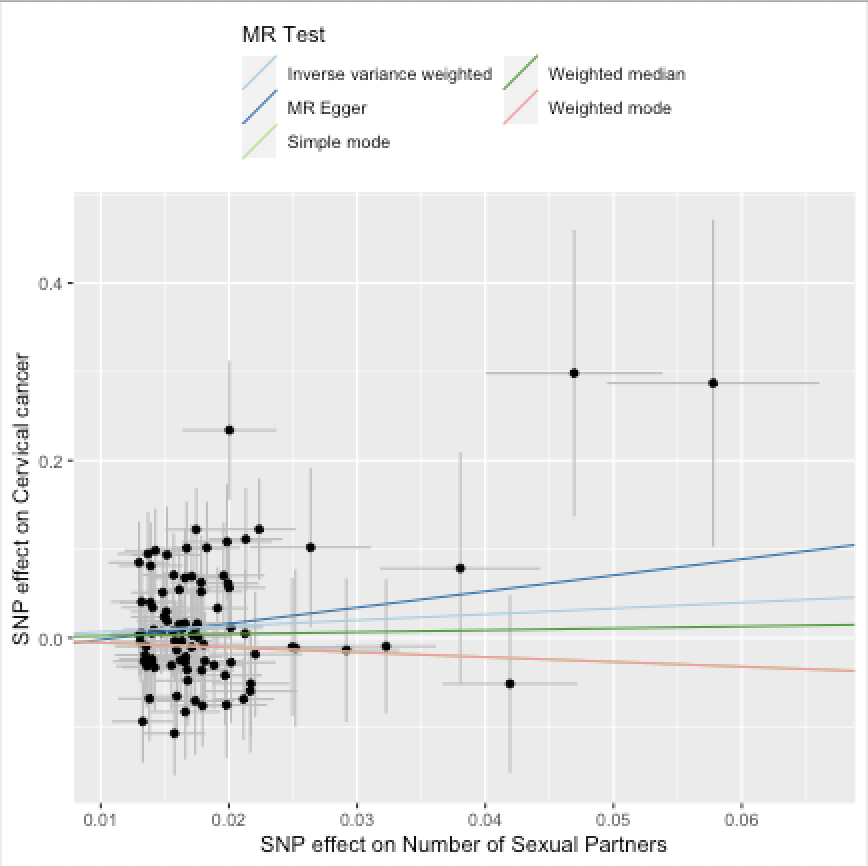

**Supplementary Fig.5** Scatter and leave one out plots for age at first sex and number of sexual partners single nucleotide polymorphisms effect on risk of C. trachomatis seropositivity

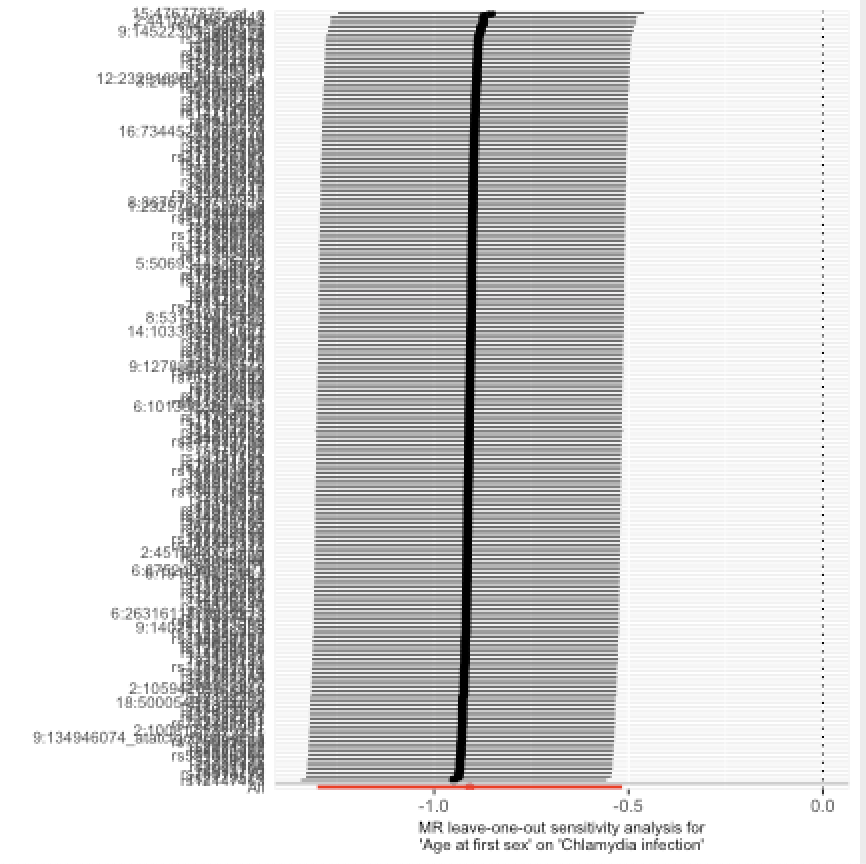

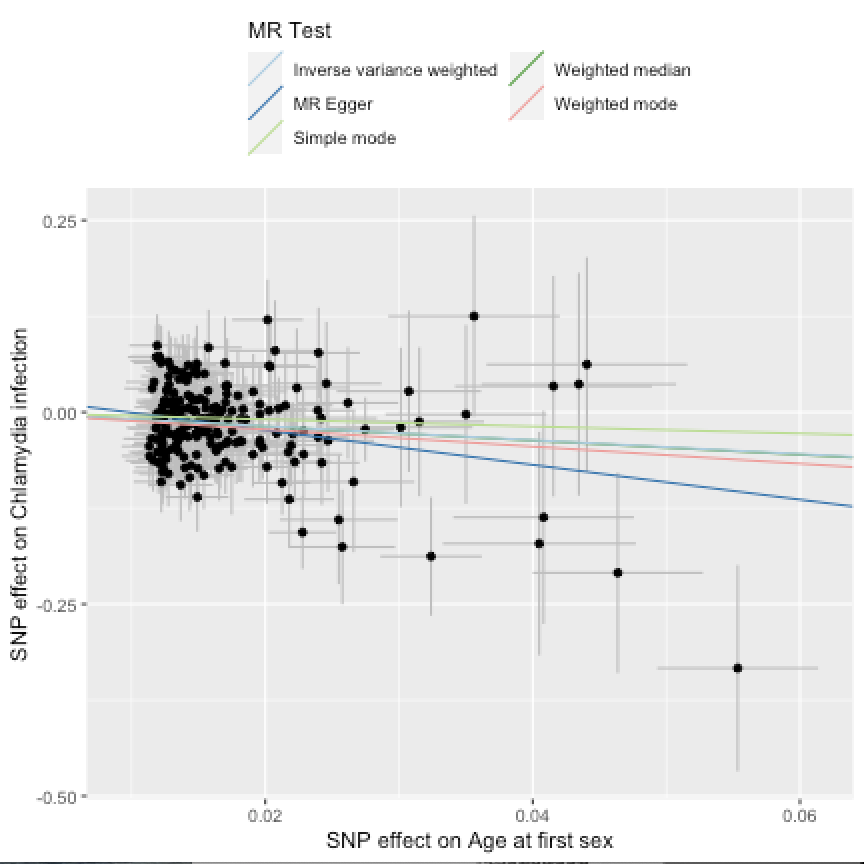

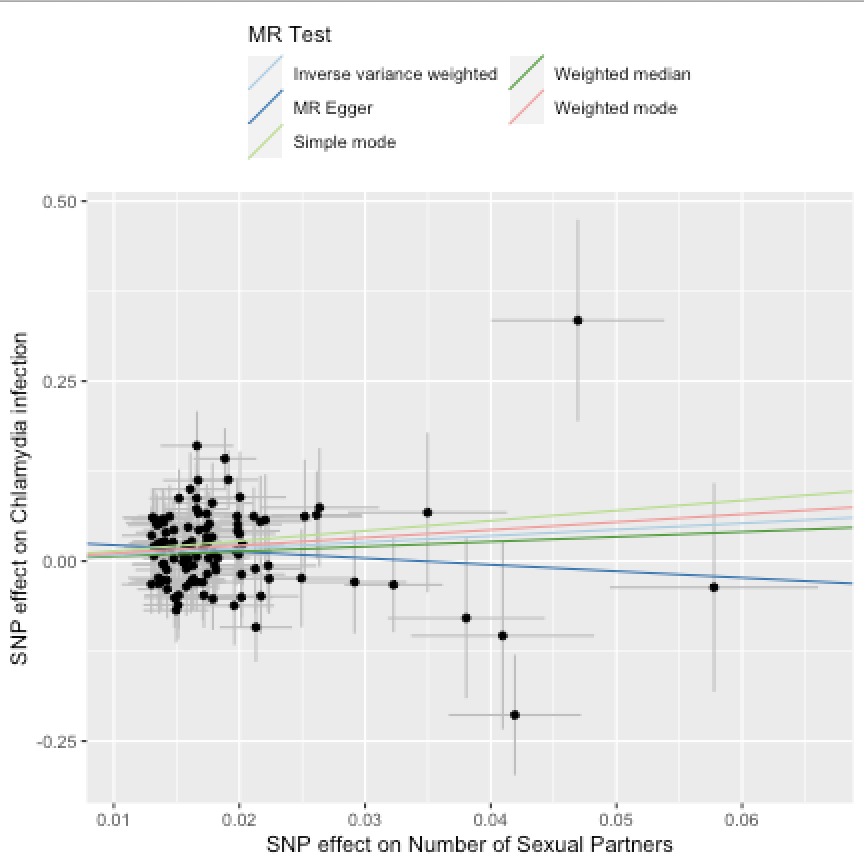

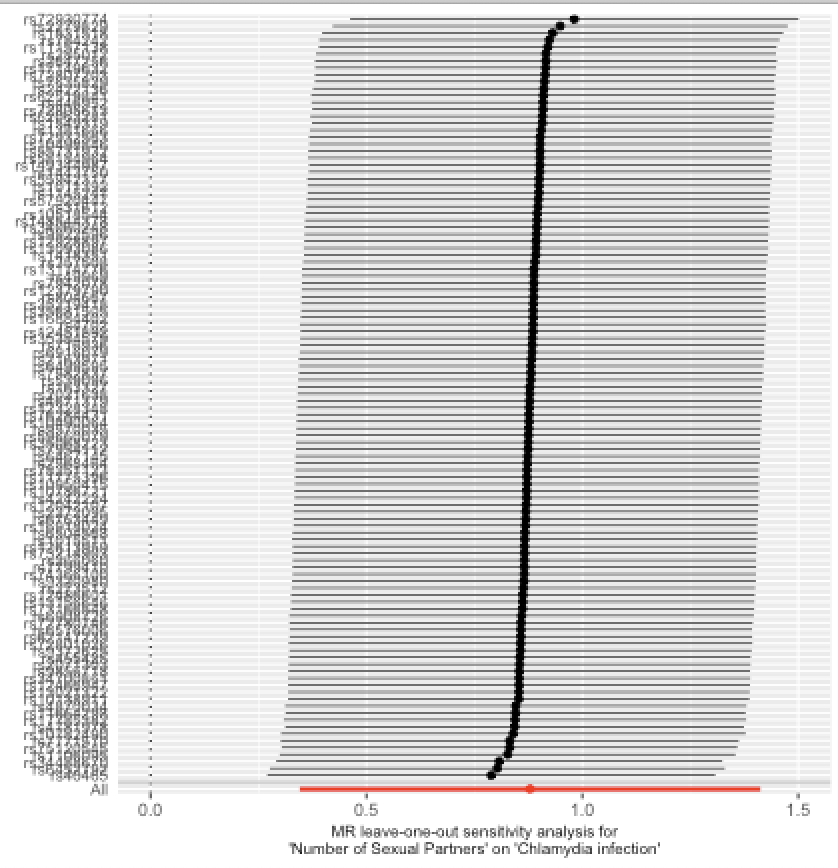

**Supplementary Fig.6** Scatter and leave one out plots for age at first sex and number of sexual partners single nucleotide polymorphisms effect on risk of lung cancer

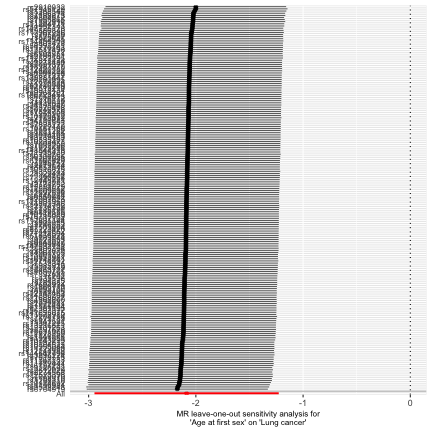

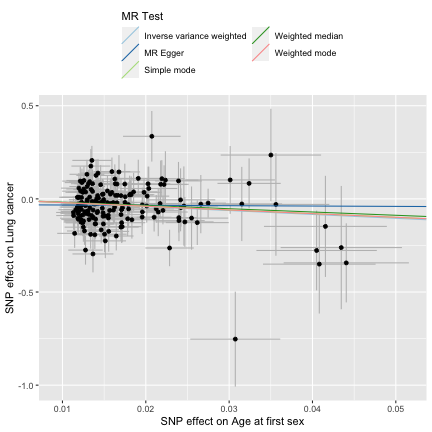

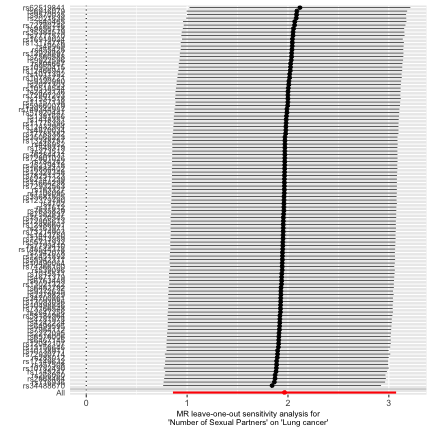

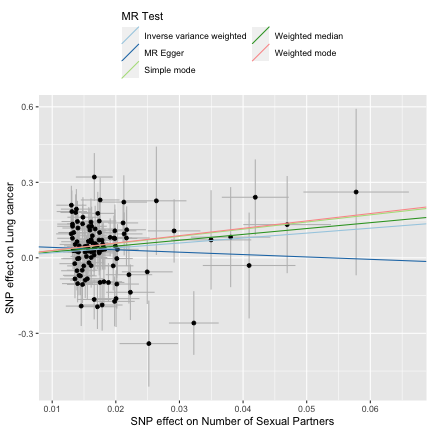

**Supplementary Fig.7** Scatter and leave one out plots for age at first sex and number of sexual partners single nucleotide polymorphisms effect on risk of oral cancer

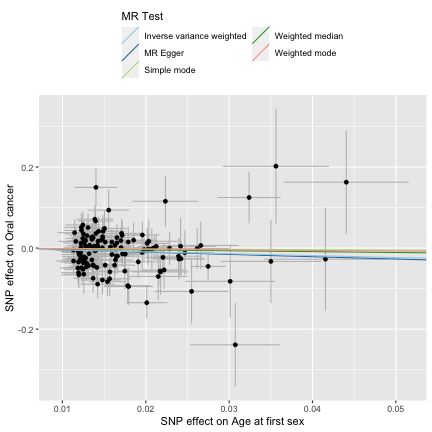

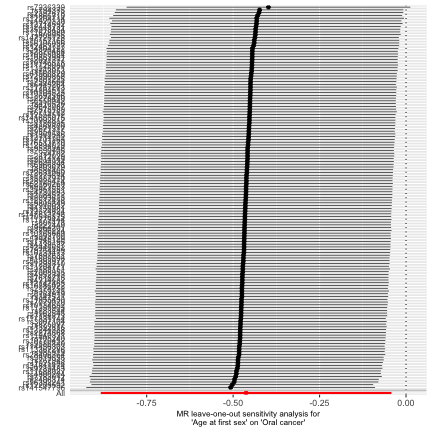

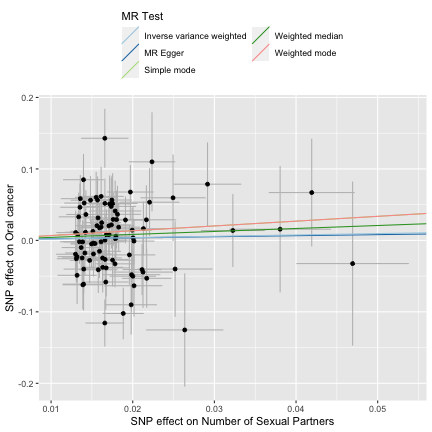

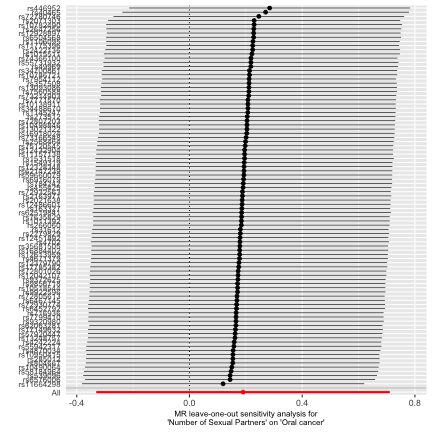

**Supplementary Fig.8** Causal Analysis Using Summary Effect estimates (CAUSE) results for age at first sex on oropharyngeal cancer

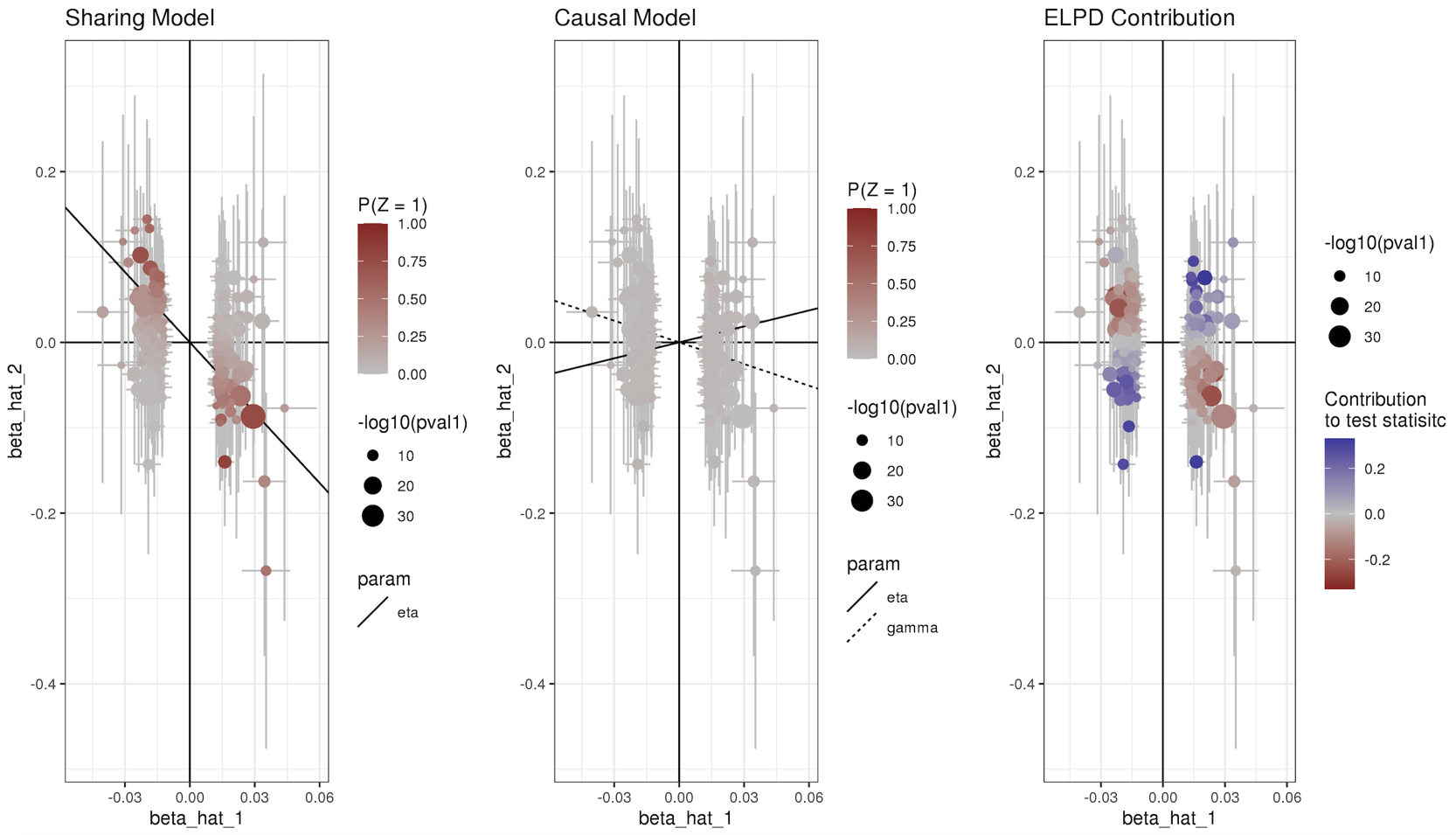

**Supplementary Fig.9** Causal Analysis Using Summary Effect estimates (CAUSE) results for number of sexual partners on oropharyngeal cancer

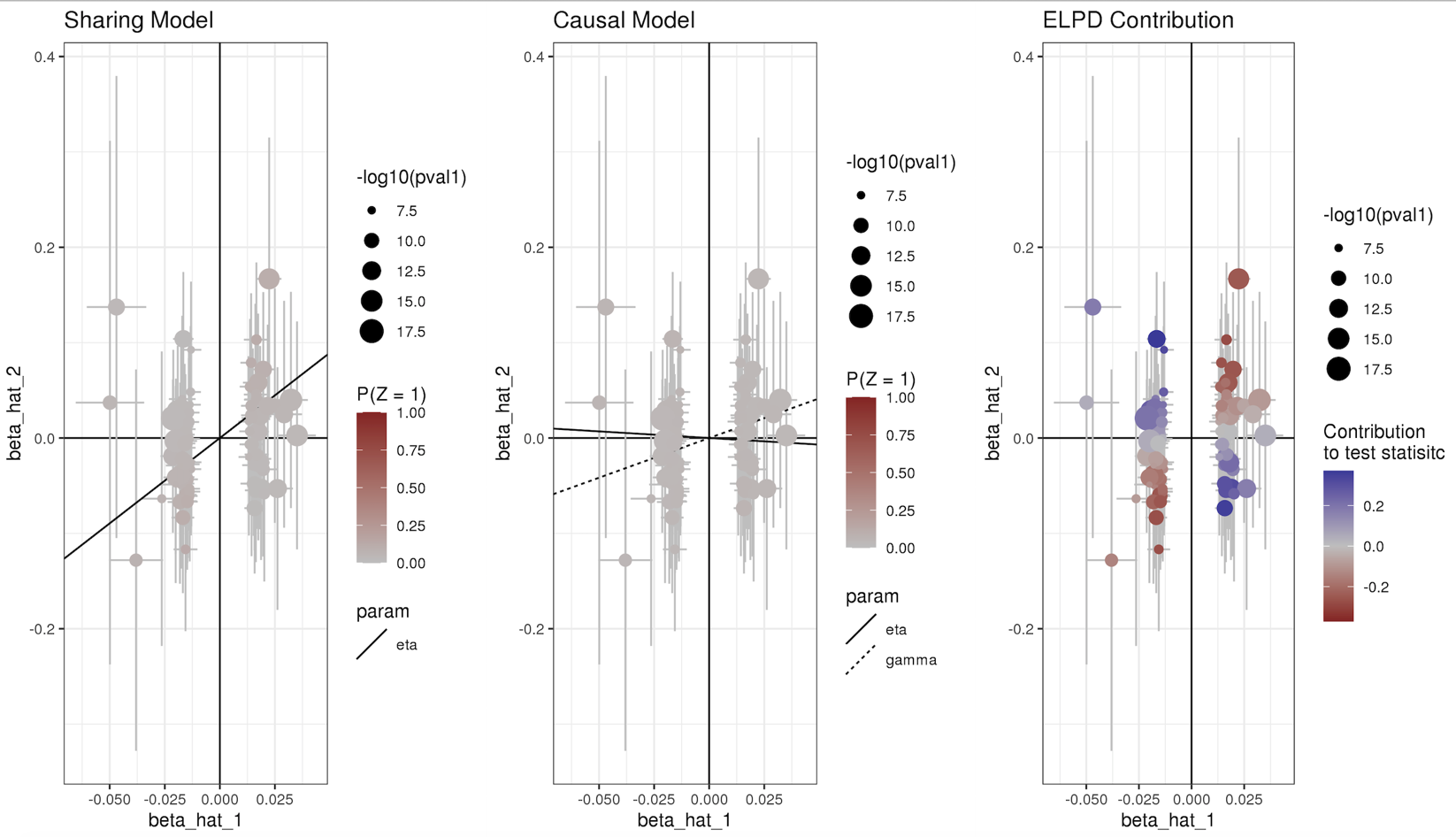

**Supplementary Fig.10** Heat map of LD Score Regression results for all exposures.

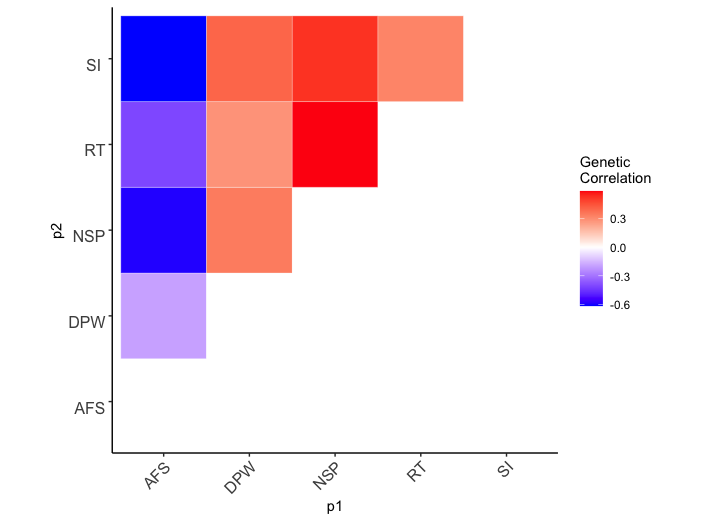

Abbreviations: AFS, age at first sex; NSP, number of sexual partners; CSI, comprehensive smoking index; SI, smoking initiation; DPW, drinks per week; RT, risk tolerance.

**Supplementary Fig.11** Forest plot showing multivariable Mendelian randomization results for age at first sex and number of sexual partners single nucleotide polymorphisms with risk of lung cancer.

**A**

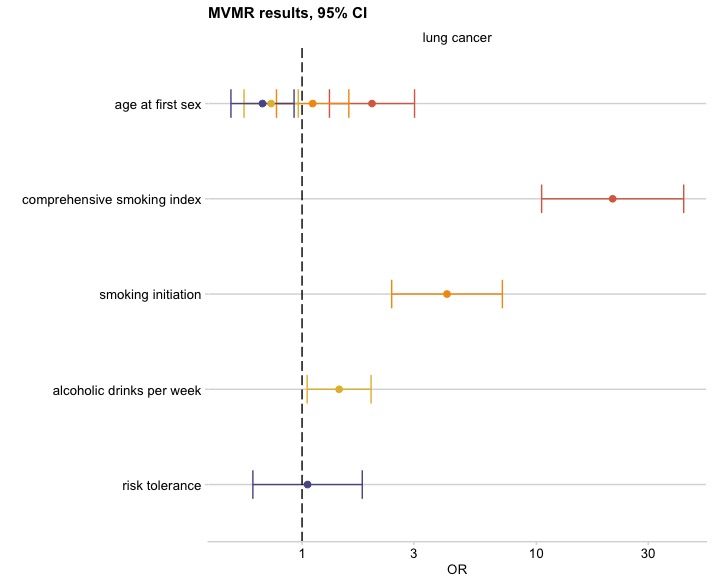

**B**

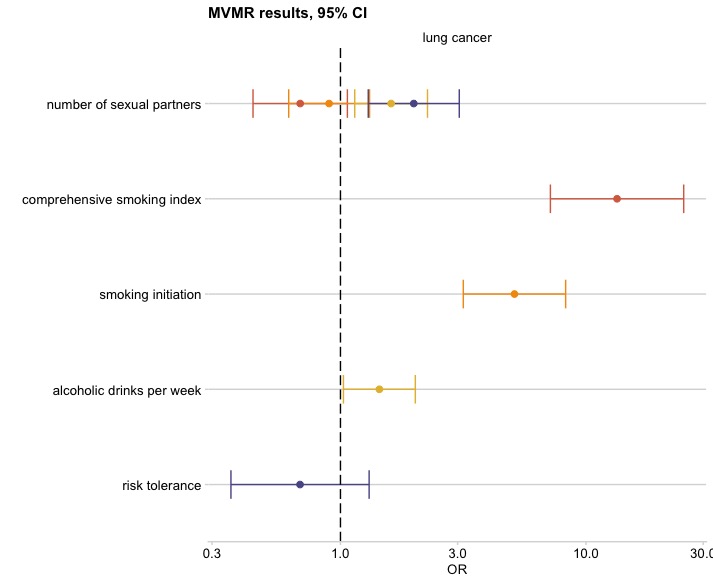

Effect estimates on oropharyngeal cancer risk are reported on the log odds scale with 95% confidence intervals. **A** Age at first sex OR represents the change in odds of lung cancer per SD change (7.3-month delay) in age at first sex. **B** Number of sexual partners OR represents the change in odds of lung cancer per SD change (0.94) in number of sexual partners. Comprehensive smoking index (dark orange), smoking initiation (light orange), alcoholic drinks per week (yellow), risk tolerance (blue).

**References**

1	Bowden, J., Smith, G. D., Haycock, P. C. & Burgess, S. Consistent Estimation in Mendelian Randomization with Some Invalid Instruments Using a Weighted Median Estimator. *Genet Epidemiol* **40**, 304-314 (2016).

2	Hartwig, F. P., Smith, G. D. & Bowden, J. Robust inference in summary data Mendelian randomization via the zero modal pleiotropy assumption. *Int J Epidemiol* **46**, 1985-1998 (2017).

3	Bowden, J., Davey Smith, G. & Burgess, S. Mendelian randomization with invalid instruments: effect estimation and bias detection through Egger regression. *Int J Epidemiol* **44**, 512-525 (2015).

4	Bowden, J. *et al.* Assessing the suitability of summary data for two-sample Mendelian randomization analyses using MR-Egger regression: the role of the I-2 statistic. *Int J Epidemiol* **45**, 1961-1974 (2016).

5	Verbanck, M., Chen, C. Y., Neale, B. & Do, R. Publisher Correction: Detection of widespread horizontal pleiotropy in causal relationships inferred from Mendelian randomization between complex traits and diseases. *Nat Genet* **50**, 1196 (2018).

6	Mentzer, A. J. *et al.* Identification of host-pathogen-disease relationships using a scalable Multiplex Serology platform in UK Biobank. *medRxiv*, 19004960 (2019).

7	Bycroft, C. *et al.* Genome-wide genetic data on ~500,000 UK Biobank participants. *bioRxiv*, 166298 (2017).

8	Waterboer, T. *et al.* Multiplex Human Papillomavirus Serology Based on In Situ–Purified Glutathione S-Transferase Fusion Proteins. *Clinical Chemistry* **51**, 1845-1853 (2005).

9	Waterboer, T., Sehr, P. & Pawlita, M. Suppression of non-specific binding in serological Luminex assays. *Journal of Immunological Methods* **309**, 200-204 (2006).

10	Kreimer, A. R. *et al.* Kinetics of the Human Papillomavirus Type 16 E6 Antibody Response Prior to Oropharyngeal Cancer. *JNCI: Journal of the National Cancer Institute* **109** (2017).

11	Hammer, C. *et al.* Amino Acid Variation in HLA Class II Proteins Is a Major Determinant of Humoral Response to Common Viruses. *Am J Hum Genet* **97**, 738-743 (2015).

12	Brenner, N. *et al.* Characterization of human papillomavirus (HPV) 16 E6 seropositive individuals without HPV-associated malignancies after 10 years of follow-up in the UK Biobank. *EBioMedicine* **62**, 103123 (2020).

13	Chang, C. C. *et al.* Second-generation PLINK: rising to the challenge of larger and richer datasets. *Gigascience* **4**, 7 (2015).

14	Trabert, B. *et al.* Antibodies Against Chlamydia trachomatis and Ovarian Cancer Risk in Two Independent Populations. *J Natl Cancer Inst* **111**, 129-136 (2019).

15	Horner, P. J. *et al.* Chlamydia trachomatis Pgp3 Antibody Persists and Correlates with Self-Reported Infection and Behavioural Risks in a Blinded Cohort Study. *PLoS One* **11**, e0151497 (2016).

16	Graff, R. E. *et al.* Cross-cancer evaluation of polygenic risk scores for 16 cancer types in two large cohorts. *Nat Commun* **12**, 970 (2021).

17	Wang, Y. *et al.* Rare variants of large effect in BRCA2 and CHEK2 affect risk of lung cancer. *Nat Genet* **46**, 736-741 (2014).

18	Lesseur, C. *et al.* Genome-wide association analyses identify new susceptibility loci for oral cavity and pharyngeal cancer. *Nat Genet* **48**, 1544-1550 (2016).

19	Dudding, T. *et al.* Assessing the causal association between 25-hydroxyvitamin D and the risk of oral and oropharyngeal cancer using Mendelian randomization. *Int J Cancer* **143**, 1029-1036 (2018).

20	Mills, M. C. *et al.* Identification of 370 loci for age at onset of sexual and reproductive behaviour, highlighting common aetiology with reproductive biology, externalizing behaviour and longevity. *bioRxiv*, 2020.2005.2006.081273 (2020).

21	Karlsson Linner, R. *et al.* Genome-wide association analyses of risk tolerance and risky behaviors in over 1 million individuals identify hundreds of loci and shared genetic influences. *Nat Genet* **51**, 245-257 (2019).

22	Wootton, R. E. *et al.* Evidence for causal effects of lifetime smoking on risk for depression and schizophrenia: a Mendelian randomisation study. *Psychol Med*, 1-9 (2019).

23	Liu, M. Z. *et al.* Association studies of up to 1.2 million individuals yield new insights into the genetic etiology of tobacco and alcohol use. *Nat Genet* **51**, 237 (2019).
