## Supplementary material for "Investigating the effect of sexual behaviour on oropharyngeal cancer risk: a methodological assessment of Mendelian randomization": Conflict of interest

**ICMJE DISCLOSURE FORM**

**Date:** 21/06/2021

**Your Name:** Mark Gormley

**Manuscript number (if known):** NA

**In the interest of transparency, we ask you to disclose all relationships/activities/interests listed below that are**

**related to the content of your manuscript. “Related” means any relation with for-profit or not-for-profit third**

**parties whose interests may be affected by the content of the manuscript. Disclosure represents a commitment**

**to transparency and does not necessarily indicate a bias. If you are in doubt about whether to list a relationship/activity/interest, it is preferable that you do so.**

**The following questions apply to the** **author’s relationships/activities/interests as they relate to the current**

**manuscript only.**

**The author’s relationships/activities/interests should be defined broadly. For example, if your manuscript pertains**

**to the epidemiology of hypertension, you should declare all relationships with manufacturers of antihypertensive medication, even if that medication is not mentioned in the manuscript.**

**In item #1 below, report all support for the work reported in this manuscript without time limit. For all other items,**

**the time frame for disclosure is the past 36 months.**

|  |  | **Name all entities with whom you have this relationship or indicate none (add rows as needed)** | | **Specifications/Comments**  **(e.g., if payments were made to you or to your institution)** |
| --- | --- | --- | --- | --- |
| **Time frame: Since the initial planning of the work** | | | | |
| 1 | All support for the present manuscript (e.g., funding, provision of study materials, medical writing, article processing charges, etc.)  **No time limit for this item.** | ____ None | | M.G. was a National Institute for Health Research (NIHR) academic clinical fellow and is currently supported by a Wellcome Trust GW4-Clinical Academic Training PhD Fellowship. This research was funded in part, by the Wellcome Trust [Grant number 220530/Z/20/Z]. For the purpose of open access, the author has applied a CC BY public copyright licence to any Author Accepted Manuscript version arising from this submission. R.C.R. is a de Pass VC research fellow at the University of Bristol. J.T. is supported by an Academy of Medical Sciences (AMS) Springboard award, which is supported by the AMS, the Wellcome Trust, Global Challenges Research Fund (GCRF), the Government Department of Business, Energy and Industrial strategy, the British Heart Foundation and Diabetes UK (SBF004\1079). R.M.M. was supported by a Cancer Research UK (C18281/A20919) programme grant (the Integrative Cancer Epidemiology Programme). R.M.M. and A.R.N. are supported by the National Institute for Health Research (NIHR) Bristol Biomedical Research Centre which is funded by the National Institute for Health Research (NIHR) and is a partnership between University Hospitals Bristol NHS Foundation Trust and the University of Bristol. Department of Health and Social Care disclaimer: The views expressed are those of the authors and not necessarily those of the NHS, the NIHR or the Department of Health and Social Care. This publication presents data from the Head and Neck 5000 study. The study was a component of independent research funded by the National Institute for Health Research (NIHR) under its Programme Grants for Applied Research scheme (RP-PG-0707-10034). The views expressed in this publication are those of the author(s) and not necessarily those of the NHS, the NIHR or the Department of Health. Core funding was also provided through awards from Above and Beyond, University Hospitals Bristol and Weston Research Capability Funding and the NIHR Senior Investigator award to A.R.N. Human papillomavirus (HPV) serology was supported by a Cancer Research UK Programme Grant, the Integrative Cancer Epidemiology Programme (C18281/A20919). B.D. and the University of Pittsburgh head and neck cancer case-control study are supported by US National Institutes of Health (NIH) grants: P50 CA097190, P30 CA047904 and R01 DE025712. The genotyping of the HNSCC cases and controls was performed at the Center for Inherited Disease Research (CIDR) and funded by the US National Institute of Dental and Craniofacial Research (NIDCR; 1X01HG007780-0). The University of North Carolina (UNC) CHANCE study was supported in part by the National Cancer Institute (R01-CA90731). E.E.V is supported by Diabetes UK (17/0005587). E.E.V is also supported by the World Cancer Research Fund (WCRF UK), as part of the World Cancer Research Fund International grant programme (IIG_2019_2009). E.H.T and P.S. were supported by FAPESP grant 10/51168-0 (GENCAPO/Head and Neck Genome project). M.G., T.D., K.B., A.C., R.M.M., M.M., G.D.S, E.E.V. and R.C.R are part of the Medical Research Council Integrative Epidemiology Unit at the University of Bristol supported by the Medical Research Council (MC_UU_00011/1, MC_UU_00011/5, MC_UU_00011/6, MC_UU_00011/7). |
|  |  | National Institute for Health Research (NIHR) | |  |
|  |  | Wellcome Trust | |  |
|  |  | University of Bristol | |  |
|  |  | Academy of Medical Sciences (AMS) | |  |
|  |  | Global Challenges Research Fund (GCRF) | |  |
|  |  | Diabetes UK | |  |
|  |  | Cancer Research UK  US National Institutes of Health (NIH)  US National Institute of Dental and Craniofacial Research  US National Cancer Institute  World Cancer Research Fund  GENCAPO  Medical Research Council (MRC) | |  |
| **Time frame: past 36 months** | | | | |
| 2 | Grants or contracts from any entity (if not indicated in item #1 above). | As above |  | |
| 3 | Royalties or licenses | __X__ None |  | |
| 4 | Consulting fees | __X__ None |  | |
| 5 | Payment or honoraria for lectures, presentations, speakers bureaus, manuscript writing or educational events | __X__ None |  | |
| 6 | Payment for expert testimony | __X__ None |  | |
| 7 | Support for attending meetings and/or travel | __X__ None |  | |
| 8 | Patents planned, issued or pending | __X__ None |  | |
| 9 | Participation on a Data  Safety Monitoring Board or Advisory Board | __X__ None |  | |
| 10 | Leadership or fiduciary role in other board, society, committee or advocacy group, paid or unpaid | __X__ None |  | |
| 11 | Stock or stock options | __X__ None |  | |
| 12 | Receipt of equipment, materials, drugs, medical writing, gifts or other services | __X__ None |  | |
| 13 | Other financial or non-financial interests | __X__ None |  | |

**Please place an “X” next to the following statement to indicate your agreement:**

**_**X**_ I certify that I have answered every question and have not altered the wording of any of the questions on this**

**form.**
